# Cell-type Specific Expression Quantitative Trait Loci Associated with Alzheimer Disease in Blood and Brain Tissue

**DOI:** 10.1101/2020.11.23.20237008

**Authors:** Devanshi Patel, Xiaoling Zhang, John J. Farrell, Jaeyoon Chung, Thor D. Stein, Kathryn L. Lunetta, Lindsay A. Farrer

## Abstract

Because regulation of gene expression is heritable and context-dependent, we investigated AD-related gene expression patterns in cell-types in blood and brain. Cis-expression quantitative trait locus (eQTL) mapping was performed genome-wide in blood from 5,257 Framingham Heart Study (FHS) participants and in brain donated by 475 Religious Orders Study/Memory & Aging Project (ROSMAP) participants. The association of gene expression with genotypes for all cis SNPs within 1Mb of genes was evaluated using linear regression models for unrelated subjects and linear mixed models for related subjects. Cell type-specific eQTL (ct-eQTL) models included an interaction term for expression of “proxy” genes that discriminate particular cell type. Ct-eQTL analysis identified 11,649 and 2,533 additional significant gene-SNP eQTL pairs in brain and blood, respectively, that were not detected in generic eQTL analysis. Of note, 386 unique target eGenes of significant eQTLs shared between blood and brain were enriched in apoptosis and Wnt signaling pathways. Five of these shared genes are established AD loci. The potential importance and relevance to AD of significant results in myeloid cell-types is supported by the observation that a large portion of GWS ct-eQTLs map within 1Mb of established AD loci and 58% (23/40) of the most significant eGenes in these eQTLs have previously been implicated in AD. This study identified cell-type specific expression patterns for established and potentially novel AD genes, found additional evidence for the role of myeloid cells in AD risk, and discovered potential novel blood and brain AD biomarkers that highlight the importance of cell-type specific analysis.

## INTRODUCTION

Recent expression quantitative trait locus (eQTL) analysis studies suggest that changes in gene expression have a role in the pathogenesis of AD ^1, 2^. However, regulation of gene expression, as well as many biological functions, has been shown to be context-specific (e.g., tissue and cell-types, developmental time point, sex, disease status, and response to treatment or stimulus) ^3-6^. One study of 500 healthy subjects found over-representation of T cell-specific eQTLs in susceptibility alleles for autoimmune disease and AD risk alleles polarized for monocyte-specific eQTL effects ^7^. In addition, disease and trait-associated cis-eQTLs were more cell type specific than average cis-eQTLs ^7^. Another study classified 12% of more than 23000 eQTLs in blood as cell-type specific ^5^. Large eQTL studies across multiple human tissues have been conducted by the GTEx consortium, with a study on genetic effects on gene expression levels across 44 human tissues collected from the same donors characterizing patterns of tissue specificity recently published ^8^.

Microglia, monocytes and macrophages share a similar developmental lineage and are all considered to be myeloid cells ^9^. It is believed that a large proportion of AD genetic risk can be explained by genes expressed in myeloid cells and not other cell-types ^10^. Several established AD genes are highly expressed in microglia ^9, 11^ and a variant in the AD-associated locus *CELF1* has been associated with lower expression of *SPI1* in monocytes and macrophages ^10^. AD risk alleles have been shown to be enriched in myeloid specific epigenomic annotations and in active enhancers of monocytes, macrophages, and microglia ^12^, and to be polarized for cis-eQTL effects in monocytes ^7^. These findings suggest that a cell-type specific analysis in blood and brain tissue may identify novel and more precise AD associations that may help elucidate regulatory networks. In this study, we performed a genome-wide *cis* ct-eQTL analysis in blood and brain, respectively, then compared eQTLs and cell-type specific eQTLs (ct-eQTLs) between brain and blood with a focus on genes, loci, and cell-types previously implicated in AD risk by genetic approaches.

## MATERIALS, SUBJECTS AND METHODS

### Study cohorts

#### Framingham Heart Study (FHS)

The FHS is a multigenerational study of health and disease in a prospectively followed community-based and primarily non-Hispanic white sample. Procedures for assessing dementia and determining AD status in this cohort are described elsewhere ^13^. Clinical, demographic, and pedigree information, as well as 1000 Genomes Project Phase 1 imputed SNP genotypes and Affymetrix Human Exon 1.0 ST array gene expression data from whole blood, were obtained from dbGaP (https://www.ncbi.nlm.nih.gov/projects/gap/cgi-bin/study.cgi?study_id=phs000007.v31.p12). Requisite information for this study was available for 5,257 participants. Characteristics of these subjects are provided in Table S1.

#### Religious Orders Study (ROS)/ Memory and Aging Project (MAP)

ROS enrolled older nuns and priests from across the US, without known dementia for longitudinal clinical analysis and brain donation and MAP enrolled older subjects without dementia from retirement homes who agreed to brain donation at the time of death ^14, 15^. RNA-sequencing brain gene expression and whole-genome sequencing (WGS) genotype data were obtained from the AMP-AD knowledge portal (https://www.synapse.org/#!Synapse:syn3219045) ^16^.

#### Data processing

Generation and initial quality control (QC) procedures of the FHS GWAS and expression data are described elsewhere and include all genotype QC and pre-adjustment of gene expression levels for batch effects and other technical covariates ^13^. ROSMAP gene expression data were log-normalized and adjusted for known and hidden variables detected by surrogate variable analysis (SVA) ^17^ in order to determine which of these variables should be included as covariates in analysis models for eQTL discovery. Additional filtering steps of FHS and ROSMAP GWAS and gene expression data included eliminating subjects with missing data, restricting gene expression data to protein coding genes, and retaining common variants (MAF□≥ 0.05) with good imputation quality (R^2^□≥□0.3).

#### Cis eQTL mapping

Cis-eQTL mapping was performed using a genome-wide design (Fig. S1). The association of gene expression with SNP genotypes for all cis SNPs within1 Mb of protein-coding genes was evaluated using linear mixed models adjusting for family structure in FHS and linear regression models for unrelated individuals in ROSMAP. In FHS, lmekin function in the R kinship package (version 1.1.3) ^18^ was applied assuming an additive genetic model with covariates for age and sex, and family structure modeled as a random-effects term for kinship - a matrix of kinship coefficients calculated from pedigree structures. The linear model for analysis of FHS can data be expressed as follows:

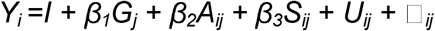

where *Y*_*i*_ is the expression value for gene *i, G*_*j*_ is the genotype dosage for cis SNP j, *Aij* and *S*_*ij*_ are the covariates for age and sex respectively, *U*_*ij*_ is the random effect for family structure, and *β*_*1*_, *β*_*2*_, and *β*_*3*_ are regression coefficients.

ROSMAP data were analyzed using the lm function in the base stats package in R ^19^. The regression model, which included covariates for age, sex, post-mortem interval (PMI), study (ROS or MAP), and a term for a surrogate variable (SV1) derived from analysis of high dimensional data, can be expressed as:

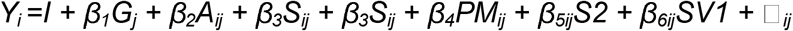

where *Y*_*i*_ is the expression value for gene *i, G*_*j*_ is the genotype dosage for cis SNP j, *Aij, S*_*ij*_, *PM*_*ij*_, *S2*_*ij*_, and *SV1*_*ij*_ are the covariates for age, sex, PMI, study and SV1 respectively, □_ij_ is the residual error, and the *β*s are regression coefficients.

#### Cis ct-eQTL mapping

Models testing associations with cell type-specific eQTLs (ct-eQTLs) included an interaction term for expression levels of “proxy” genes that represent cell types. Proxy genes representing 10 cell types in whole blood ^5^ and five cell types in brain ^20-22^ were incorporated in cell type-specific models (Table S2). These proxy genes for cell types in blood were established previously using BLUEPRINT expression data to validate cell-type-specific expression in each cell-type module ^5^ and the proxy genes for brain cell types have been incorporated in several studies ^20-22^. Cell type-specific expression analyses in blood of FHS participants were conducted using the following model:

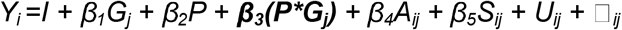

where in each eQTL_ij_ pair, *Y*_*i*_ is the eQTL expression value for gene *i, G*_*j*_ is the genotype dosage for cis SNP j, P is the proxy gene, **P\**G***_***j***_ is the interaction term representing the effect of genotype in a particular cell type, *Aij* and *S*_*ij*_ are covariates for age and sex respectively, *U*_*ij*_ is the random effect for family structure, and *β*s are regression coefficients. Models with significant interaction terms indicate cell type specific eQTLs.

The following model was used to evaluate cell type-specific expression in brain in ROSMAP:

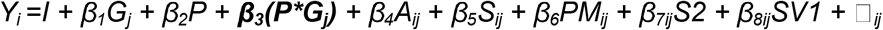

where in each eQTL_ij_ pair, variables *Y*_*i*_, *G*_*j*_, P, *Aij, S*_*ij*_, *P*_*ij*_, □_ij_ and *β*s are as described above, and *PM*_*ij*_, *S2*_*ij*_, and *SV1*_*ij*_ are covariates for PMI, study, and SV1 respectively.

A Bonferroni correction was applied to determine the significance threshold for each analysis (Table S3).

#### Selection of eQTLs in established AD loci and gene-set pathway enrichment analysis

eGenes (genes whose expression levels are associated with variation at a particular eSNP) were matched to 88 genes located near 80 distinct uncorrelated SNPs that have been associated with AD or AD-related traits by genetic association or experimental approaches (Table S4) and eSNPs (SNPs that significantly influence gene expression) under the 80 significant association peaks. Gene-set enrichment analysis was performed using the PANTHER (Protein ANalysis THrough Evolutionary Relationships) software tool ^23^ to determine if the unique genes in the significant eQTL/ct-eQTL pairs shared by both brain and blood datasets are associated with a specific biological process or molecular function. Significance of the pathways was determined by the Fisher’s Exact test with False discovery rate (FDR) multiple test correction.

#### Colocalization analyses

Assessment of causal variants shared by adjacent GWAS and eQTL signals was performed using a Bayesian colocalization approach implemented in the R package *coloc* ^24^. This analysis incorporated information about significantly associated variants for AD risk obtained from a recent large GWAS ^25^ and lead eQTL variants each defined as the eSNP showing the strongest association with gene expression. Following recommended guidelines, the variants were deemed to be colocalized by a high posterior probability that a single shared variant is responsible for both signals (PP4 > 0.8) ^24, 26^. A lower threshold for statistical significance with a false discovery rate (FDR) < 0.05 for eQTL significant results was applied to maximize detection of colocalized pairs. Regional plots were constructed with LocusZoom ^27^.

## RESULTS

A total of 847,429 eQTLs and 30,405 ct-eQTLs in blood, and 173,857 eQTLs and 51,098 ct-eQTLs in brain were significant after Bonferroni correction (Table S3 and Supplemental Resources). Among these results, 11,649 ct-eQTLs in brain 2,533 ct-eQTLs in blood involved SNP-eGene pairs that were not detected in eQTL analysis (Fig. 1A). Of note, 24,028 significant eQTLs were shared between blood and brain. The 386 distinct eGenes among these shared eQTLs (Table S5) are most enriched in the apoptosis signaling *(P=*0.023) and Wnt signaling (*P*=0.036) pathways (Table S6). Five of these eGenes (*HLA-DRB5, HLA-DRB1, ECHDC3, CR1*, and *WWOX*) were previously associated with AD ^25, 28^. Three eSNPs in eQTLs involving *HLA-DRB1*/*HLA-DRB5* (rs9271058) and *ARL17A*/*LRRC37A2* (rs2732703 and rs113986870, which are near *KANSL1* and *MAPT*) were previously associated with AD risk at the genome-wide significance level ^25, 29^.

**Figure 1.**
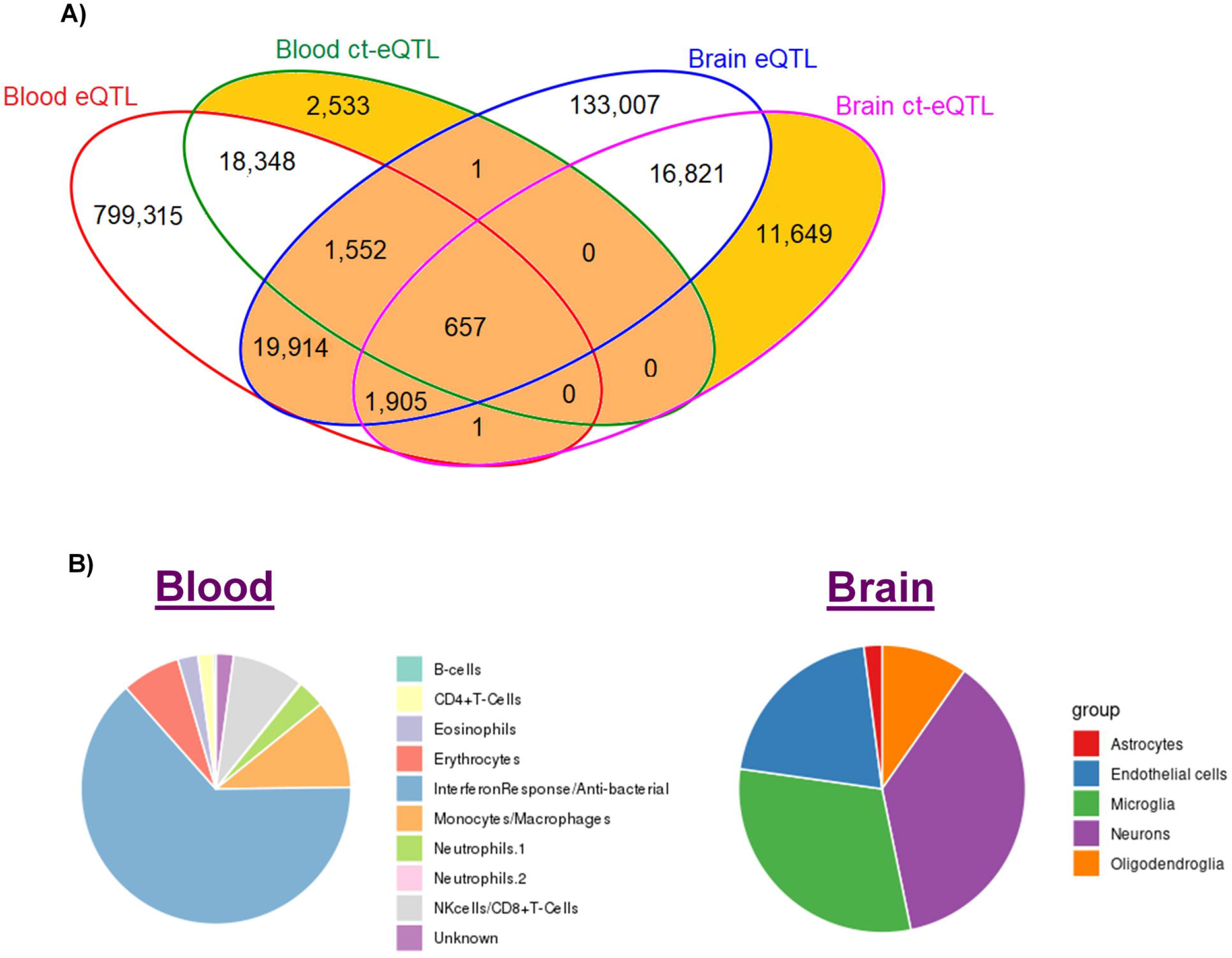
Significant gene-SNP eQTLs and ct-eQTLs in blood and brain tissue genome-wide. **A)** Venn diagram shows the number of overlapping eQTLs and ct-eQTLs in blood and brain. Gold color indicates significant eQTLs that are cell-type specific. Orange color indicates significant eQTLs that are shared between blood and brain. **B)** Cell-type distributions of significant genome-wide ct-eQTL results in blood and brain.

eQTLs involving *CR1, ECHDC3* and *WWOX* were much more significant in brain than blood, whereas *HLA-DRB5* and *HLA-DRB1* were more significant in blood when comparing the effect sizes (Table 1). *ECHDC3* was a significant eGene in blood and brain eQTLs (specifically in neurons). *HLA-DRB5* and *HLA-DRB1* were the only eGenes ascribed to significant ct-eQTLs in both blood and brain noting that of the 10 distinct lead eSNPs, five are unique to each tissue. Although the eQTLs involving these genes with the largest effect were observed in blood across multiple cell types, the total number of significant eSNP-eGene combinations was far greater in brain (particularly in microglia and neurons). The only instance in which the lead eSNP is also associated with AD risk at the GWS level was observed in the blood eQTL pair of *HLA-DRB1* with eSNP rs9271058 (Table 1A). Among the AD-associated SNPs at the GWS level, rs9271058 is a significant eSNP for *HLA-DRB1* in both blood and brain cell types (the most significant association by p-value was observed in anti-bacterial cells and microglia) and rs9271192 is a significant ct-eQTL for the gene in multiple brain cell types (Table 1). Both of these SNPs are also eSNPs for *HLA-DB5* in the brain in neurons only.

**Table 1:**
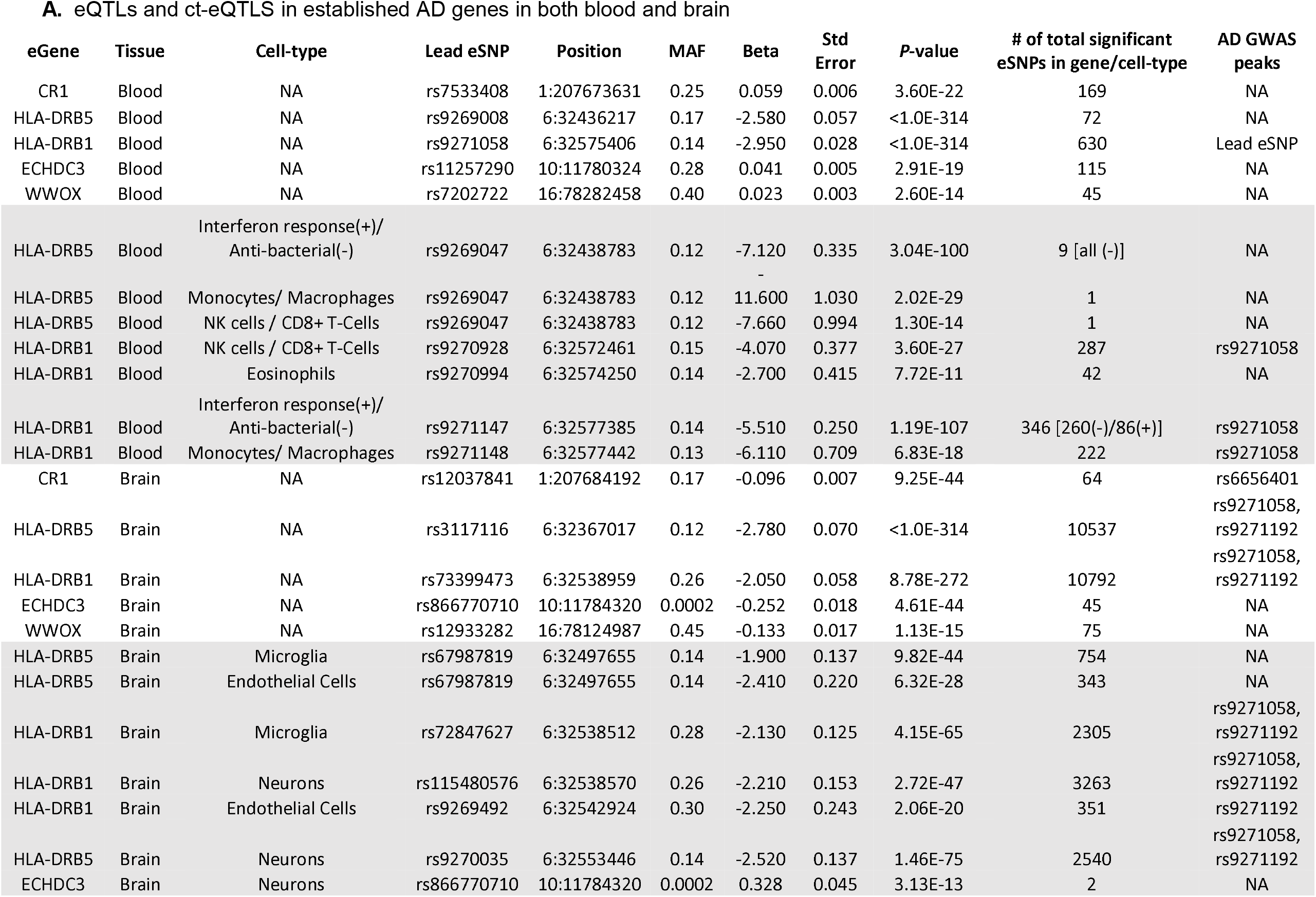
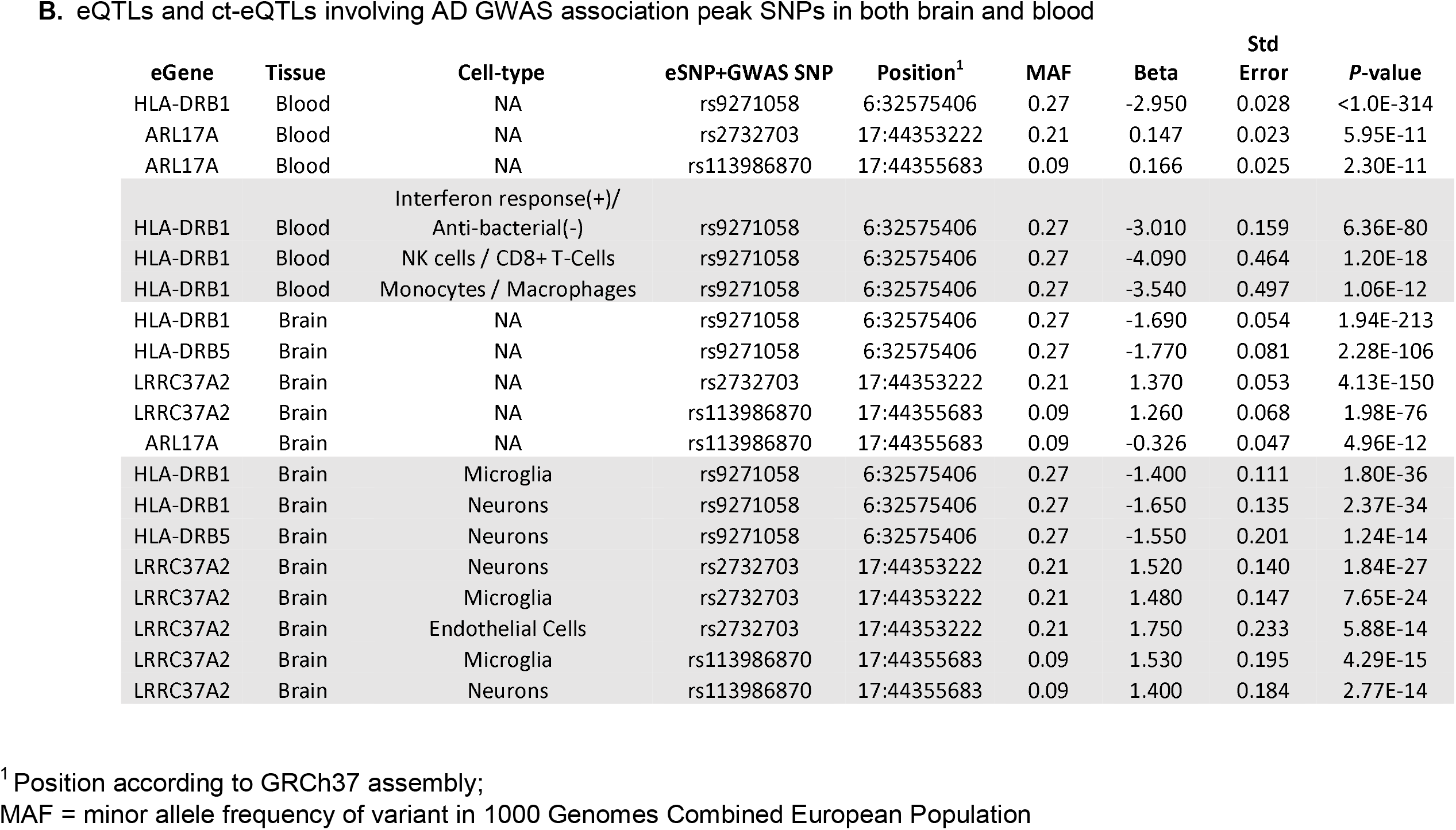
eQTLs and ct-eQTLs in established AD loci appearing in both blood and brain

There were 657 gene-SNP eQTL pairs comprising 16 unique eGenes that were significant in blood and brain overall as well as in specific cell types in both blood and brain (Table S7). None of these eGenes were observed in significant pathways enriched for AD genes, however, they included AD-associated genes *HLA-DRB1* and *HLA-DRB5*.

Slightly more than half (42/80 = 52.5%) of the established AD associations (Table S3) are eGene targets for significant eQTLs in blood (Table S8). By comparison, only seven established AD loci were eGene targets for significant eQTLs in brain, among which *OARD1* was significant in endothelial cells only (Table S8). Many GWS SNPs for AD risk are eSNPs affecting expression of the nearest gene, which is usually recognized as the causative gene, but several GWS SNPs target other genes (Table S9). For example, AD-associated eSNPs rs113986870 and rs2732703 in the *MAPT/KANSL1* region target *ARL17A* in blood, but are paired in seven of eight eQTLs and ct-eQTLs with *LRRC37A2* in brain (Table S9). *HLA-DRB1* is the only AD gene with a significant ct-eQTL in blood, whereas many AD genes have significant blood eQTLs. In brain, only four AD loci (*CR1, HLA-DRB1/DRB5, IQCK* and *MAPT/KANSL1*) have significant brain eQTLs of which *HLA-DRB1/DRB5* and *MAPT/KANSL1* are the only brain ct-eQTLs, noting that all are significant in microglia, neurons and endothelial cells.

Next, we evaluated whether the most significant eSNPs and SNPs genome-wide significantly associated with AD status (i.e. AD-SNPs) co-localize and thus to identify a single shared variant responsible for both signals (posterior probability of shared signals (PP4) > 0.8). This analysis revealed eight eQTL/ct-eQTL signals that colocalized with seven AD GWAS signals and half of the co-localized signals involved a ct-eQTL (Table 2 and Fig. S2). Two different eSNPs for *CD2AP*, rs4711880 (eQTL *P*=1.4×10^−104^) and rs13201473 (ct-eQTL *P*= 1.47×10^−9^), flank *CD2AP* GWAS SNP rs10948363 which is also the second most significant eQTL (*P*=2.32×10^−104^) and the second most significant ct-eQTL in NK cells / CD8+ T-Cells (*P*=2.66×10^−9^). These three SNPs span a 9.0 kb region in intron 2 and are in complete linkage disequilibrium (LD, r^2^□=1.0), indicating that any one or more of them could affect the function of target gene CD2AP. Rs6557994 is the most significant eSNP for and located in *PTK2B* (blood ct-eQTL *P*=2.58×10^−9^) and is moderately correlated with the *PTK2B* GWAS SNP (rs28834970, r^2^=0.78, *P*=1.58×10^−9^). Thus, it is not surprising that rs6557994 is also significantly associated with AD risk (*P*=8.19×10^−7^). Rs6557994 is also correlated with a GWAS SNP in *CLU*, located approximately 150 kb from PTK2B, that is not significantly associated with expression of any gene. Because *PTK2B* and *CLU* are independent AD risk loci ^28^, it is possible that this eSNP has an effect on AD pathogenesis through independent pathways (Fig. S2). The most significant eSNP in *MADD* (rs35233100, *P*=2.88×10^−10^) was predicted to have functional consequences because it is a stop-gained mutation. This brain eQTL is colocalized (PP4=0.95) and weakly correlated with a GWAS SNP (*P*=1.91×10^−5^) in *CELF1* rs10838725 (r^2^=0.12).

**Table 2:** Colocalized AD GWAS/lead eQTL SNP pairs

| Region <sup>1</sup> | AD GWAS Variant |  |  |  |  |  |  | Lead eQTL Variant |  |  |  |  |  | eQTL type | PP4 | r<sup>2</sup> |
| --- | --- | --- | --- | --- | --- | --- | --- | --- | --- | --- | --- | --- | --- | --- | --- | --- |
|  | rsID | Nearest Gene | MAF | p-value | eQTL P-value | eGene | Cell-type | rsID | MAF | eGene | eQTL P-value | Cell-type | GWAS P-value |  |  |  |
| 6:46487762-48487762 | rs10948363 | CD2AP | 0.72 | 1.77E-07 | 2.32E-104 | CD2AP | NA | rs4711880 | 0.23 | CD2AP | 1.36E-104 | NA | 2.57E-07 | Blood eQTL | 0.909 | 1.00 |
| 6:46487762-48487762 | rs10948363 | CD2AP | 0.72 | 1.77E-07 | 2.66E-09 | CD2AP | NK cells / CD8+ T-Cells | rs13201473 | 0.27 | CD2AP | 1.47E-09 | NK cells / CD8+ T-Cells | 2.74E-07 | Blood ct-eQTL | 0.917 | 1.00 |
| 8:26195121-28195121 | rs28834970 | PTK2B | 0.63 | 1.58E-09 | 9.15E-09 | PTK2B | Interferon response/ Anti-bacterial Cells | rs6557994 | 0.41 | PTK2B | 2.58E-09 | Interferon response/ Anti-bacterial Cells | 8.19E-07 | Blood ct-eQTL | 0.990 | 0.78 |
| 8:26467686-28467686 | rs9331896 | CLU | 0.61 | 3.62E-16 | Not an eSNP |  |  | <u>rs6557994</u> | 0.45 | PTK2B | 2.58E-09 | Interferon response/ Anti-bacterial Cells | 8.19E-07 | Blood ct-eQTL | 0.990 | 0.00 |
| 1:206692049-208692049 | rs6656401 | CR1 | 0.19 | 2.17E-15 | 1.05E-43 | CR1 | NA | rs12037841 | 0.19 | CR1 | 9.25E-44 | NA | 1.77E-15 | Brain eQTL | 0.993 | 1.00 |
| 11:46557871-48557871 | rs10838725 | CELF1 | 0.68 | 1.91E-05 | Not an eSNP |  |  | rs35233100 | 0.068 | MADD | 2.88E-10 | NA | 1.25E-03 | Brain eQTL | 0.954 | 0.12 |
| 11:58923508-60923508 | rs983392 | MS4A6A | 0.59 | 4.76E-15 | Not an eSNP |  |  | rs11230563 | 0.35 | CD6 | 2.31E-113 | NA | 0.48 | Brain eQTL | 0.854 | 0.00 |
| 19:44411941-46411941 | rs429358 | APOE | 0.78 | 0.00E+00 | Not an eSNP |  |  | rs74253343 | 0.47 | RELB | 1.9E-14 | Oligodendroglia | 0.23 | Brain ct-eQTL | 0.971 | 0.00 |

<sup>1</sup> Map position within 1 Mb of AD GWAS SNP according to GRCh37 assembly;
MAF = minor allele frequency; NA = not available; PP4 = posterior probability of colocalization; r<sup>2</sup> = correlation of AD and eQTL variants

Examination of the distribution of the significant ct-eQTL results genome-wide showed that nearly two-thirds of the ct-eQTLs in blood occurred in interferon response/anti-bacterial cells in blood, whereas brain ct-eQTLs are highly represented in endothelial cells, neurons and microglia (Fig. 1B, Table S10). Further examination of significant results within myeloid cell lineages (i.e., microglia and monocytes/ macrophages) which account for a large proportion of the genetic risk for late-onset AD ^10^ revealed that 3,234 or 10.6% of all significant ct-eQTLs in blood were in monocytes/macrophages. This subset includes 128 unique eGenes which are significantly enriched in the AD amyloid secretase pathway (FDR *P*=0.013, Table S11). A total of 974 or 30.1% of ct-eQTLs including 4 of the 20 most significant eGenes in monocytes/macrophages are located within 1 Mb of established AD loci (Table 4A). One of the eGenes in this top-ranked group (*HLA-DRB5*) is an established AD locus, and three others that are near established AD loci (*DLG2* near *PICALM* ^30^, *C4BPA* near *CR1* ^31^, and *MYO1E* near *ADAM10* ^32^) are reasonable AD gene candidates based on evidence using non-genetic approaches. Microglia accounted for 15,560 (30.5%) of significant ct-eQTLs in the brain (Table S10) which involved 304 unique eGenes. Approximately 52% of significant ct-eQTLs in microglia are located in AD regions including five of the 20 most significant ct-eQTLs in this group (Table 4B). One of these five eGenes is an established AD locus (*HLA-DRB1*) and two others (*ALCC* ^33^ and *WNT3* ^34^) have been linked to AD in previous studies.

**Table 3:**
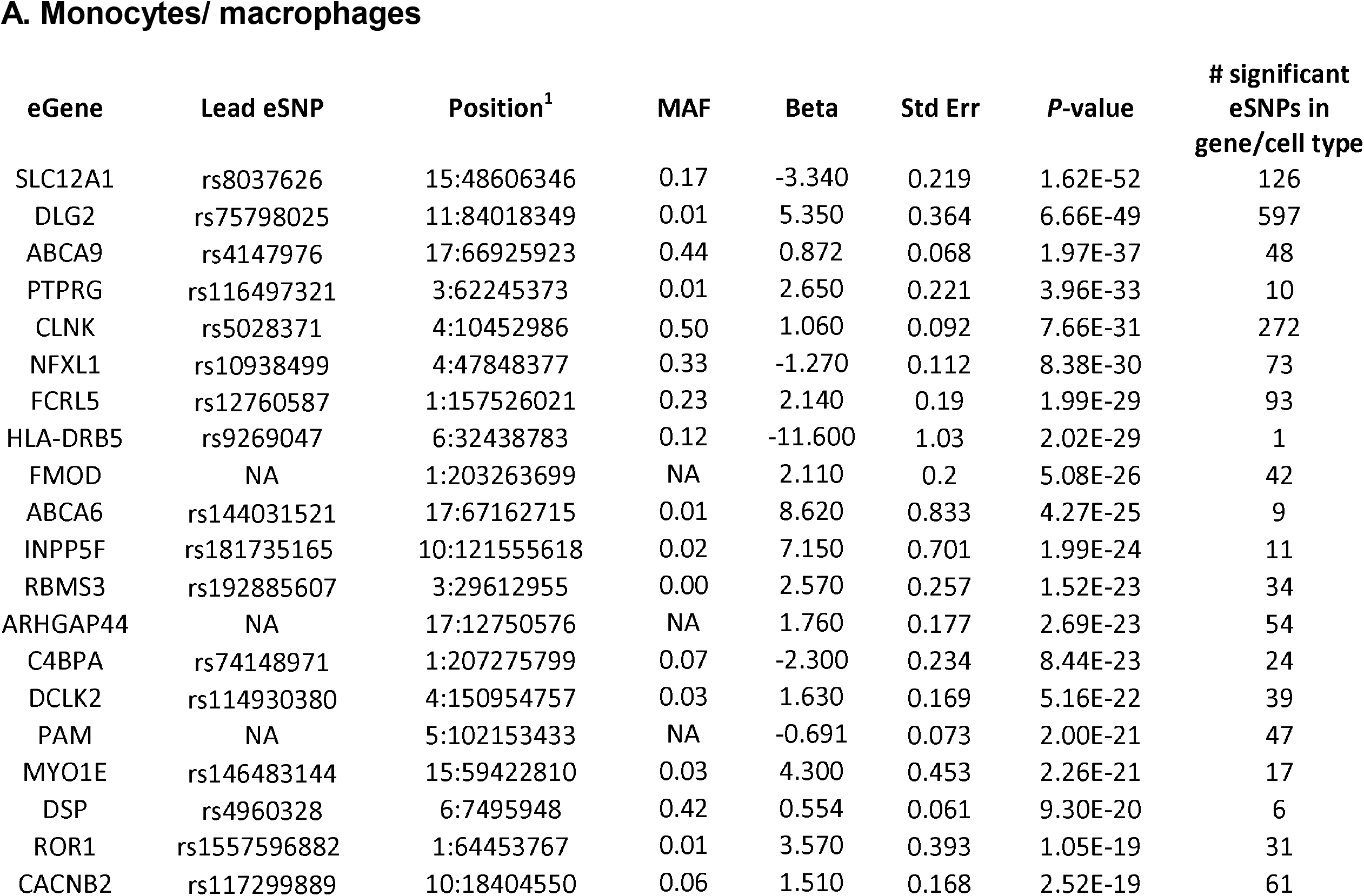
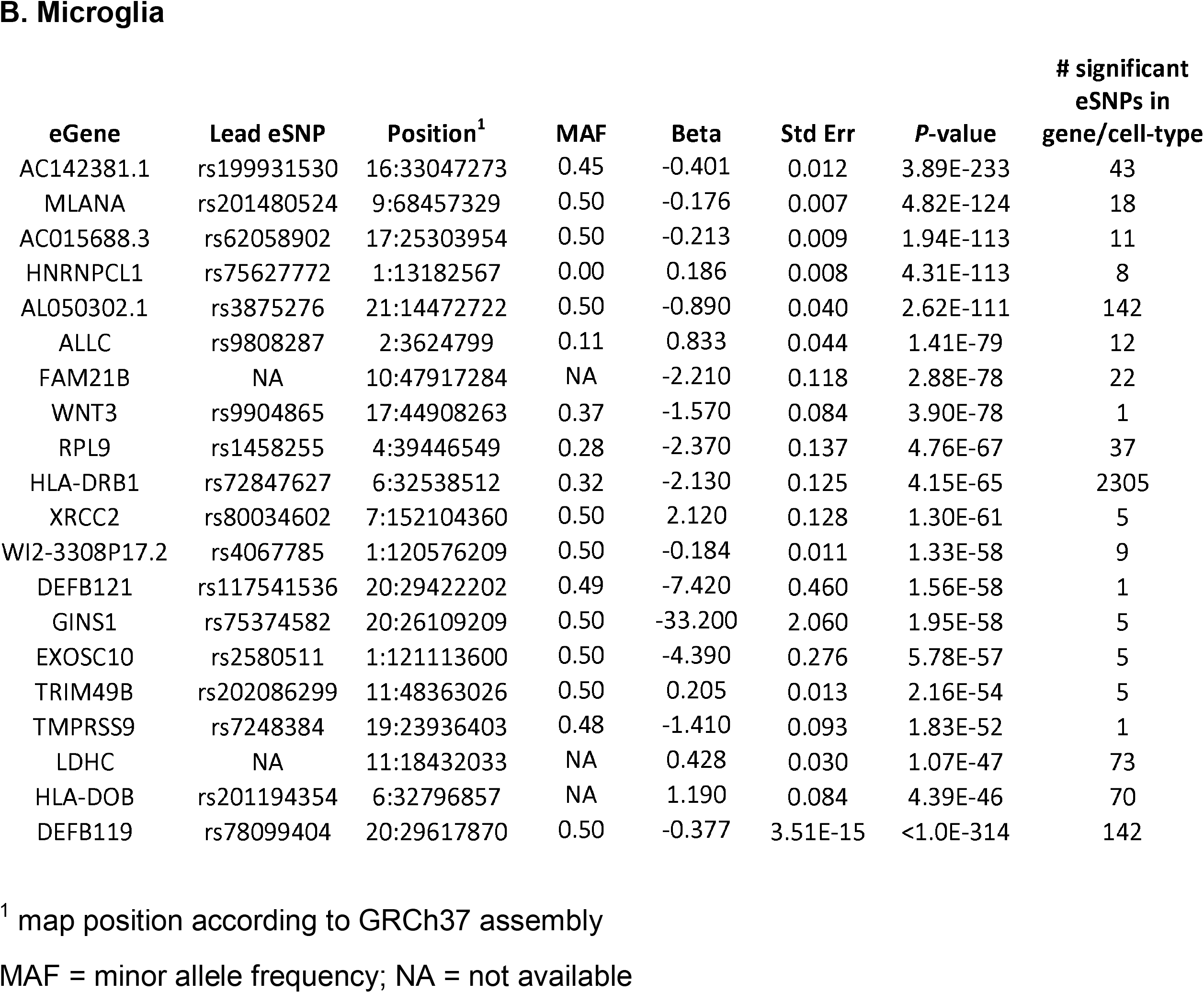
Top-ranked ct-eQTLs in myeloid cell types

| eGene | Lead eSNP | Position<sup>1</sup> | MAF | Beta | Std Err | P-value | # significant eSNPs in gene/cell type |
| --- | --- | --- | --- | --- | --- | --- | --- |
| SLC12A1 | rs8037626 | 15:48606346 | 0.17 | -3.340 | 0.219 | 1.62E-52 | 126 |
| DLG2 | rs75798025 | 11:84018349 | 0.01 | 5.350 | 0.364 | 6.66E-49 | 597 |
| ABCA9 | rs4147976 | 17:66925923 | 0.44 | 0.872 | 0.068 | 1.97E-37 | 48 |
| PTPRG | rs116497321 | 3:62245373 | 0.01 | 2.650 | 0.221 | 3.96E-33 | 10 |
| CLNK | rs5028371 | 4:10452986 | 0.50 | 1.060 | 0.092 | 7.66E-31 | 272 |
| NFXL1 | rs10938499 | 4:47848377 | 0.33 | -1.270 | 0.112 | 8.38E-30 | 73 |
| FCRL5 | rs12760587 | 1:157526021 | 0.23 | 2.140 | 0.19 | 1.99E-29 | 93 |
| HLA-DRB5 | rs9269047 | 6:32438783 | 0.12 | -11.600 | 1.03 | 2.02E-29 | 1 |
| FMOD | NA | 1:203263699 | NA | 2.110 | 0.2 | 5.08E-26 | 42 |
| ABCA6 | rs144031521 | 17:67162715 | 0.01 | 8.620 | 0.833 | 4.27E-25 | 9 |
| INPP5F | rs181735165 | 10:121555618 | 0.02 | 7.150 | 0.701 | 1.99E-24 | 11 |
| RBMS3 | rs192885607 | 3:29612955 | 0.00 | 2.570 | 0.257 | 1.52E-23 | 34 |
| ARHGAP44 | NA | 17:12750576 | NA | 1.760 | 0.177 | 2.69E-23 | 54 |
| C4BPA | rs74148971 | 1:207275799 | 0.07 | -2.300 | 0.234 | 8.44E-23 | 24 |
| DCLK2 | rs114930380 | 4:150954757 | 0.03 | 1.630 | 0.169 | 5.16E-22 | 39 |
| PAM | NA | 5:102153433 | NA | -0.691 | 0.073 | 2.00E-21 | 47 |
| MYO1E | rs146483144 | 15:59422810 | 0.03 | 4.300 | 0.453 | 2.26E-21 | 17 |
| DSP | rs4960328 | 6:7495948 | 0.42 | 0.554 | 0.061 | 9.30E-20 | 6 |
| ROR1 | rs1557596882 | 1:64453767 | 0.01 | 3.570 | 0.393 | 1.05E-19 | 31 |
| CACNB2 | rs117299889 | 10:18404550 | 0.06 | 1.510 | 0.168 | 2.52E-19 | 61 |

B. Microglia
| eGene | Lead eSNP | Position<sup>1</sup> | MAF | Beta | Std Err | P-value | # significant eSNPs in gene/cell-type |
| --- | --- | --- | --- | --- | --- | --- | --- |
| AC142381.1 | rs199931530 | 16:33047273 | 0.45 | -0.401 | 0.012 | 3.89E-233 | 43 |
| MLANA | rs201480524 | 9:68457329 | 0.50 | -0.176 | 0.007 | 4.82E-124 | 18 |
| AC015688.3 | rs62058902 | 17:25303954 | 0.50 | -0.213 | 0.009 | 1.94E-113 | 11 |
| HNRNPCL1 | rs75627772 | 1:13182567 | 0.00 | 0.186 | 0.008 | 4.31E-113 | 8 |
| AL050302.1 | rs3875276 | 21:14472722 | 0.50 | -0.890 | 0.040 | 2.62E-111 | 142 |
| ALLC | rs9808287 | 2:3624799 | 0.11 | 0.833 | 0.044 | 1.41E-79 | 12 |
| FAM21B | NA | 10:47917284 | NA | -2.210 | 0.118 | 2.88E-78 | 22 |
| WNT3 | rs9904865 | 17:44908263 | 0.37 | -1.570 | 0.084 | 3.90E-78 | 1 |
| RPL9 | rs1458255 | 4:39446549 | 0.28 | -2.370 | 0.137 | 4.76E-67 | 37 |
| HLA-DRB1 | rs72847627 | 6:32538512 | 0.32 | -2.130 | 0.125 | 4.15E-65 | 2305 |
| XRCC2 | rs80034602 | 7:152104360 | 0.50 | 2.120 | 0.128 | 1.30E-61 | 5 |
| WI2-3308P17.2 | rs4067785 | 1:120576209 | 0.50 | -0.184 | 0.011 | 1.33E-58 | 9 |
| DEFB121 | rs117541536 | 20:29422202 | 0.49 | -7.420 | 0.460 | 1.56E-58 | 1 |
| GINS1 | rs75374582 | 20:26109209 | 0.50 | -33.200 | 2.060 | 1.95E-58 | 5 |
| EXOSC10 | rs2580511 | 1:121113600 | 0.50 | -4.390 | 0.276 | 5.78E-57 | 5 |
| TRIM49B | rs202086299 | 11:48363026 | 0.50 | 0.205 | 0.013 | 2.16E-54 | 5 |
| TMPRSS9 | rs7248384 | 19:23936403 | 0.48 | -1.410 | 0.093 | 1.83E-52 | 1 |
| LDHC | NA | 11:18432033 | NA | 0.428 | 0.030 | 1.07E-47 | 73 |
| HLA-DOB | rs201194354 | 6:32796857 | NA | 1.190 | 0.084 | 4.39E-46 | 70 |
| DEFB119 | rs78099404 | 20:29617870 | 0.50 | -0.377 | 3.51E-15 | <1.0E-314 | 142 |

<sup>1</sup> map position according to GRCh37 assembly
MAF = minor allele frequency; NA = not available

**Table 4:** Overlap of ct-eQTLs in myeloid cell types in brain and blood.

|  |  |  |  |  |
| --- | --- | --- | --- | --- |
| BTNL3 | FAM118A | HLA-DOB | HLA-DRB1 | HLA-DRB5 |
| 1 | 43 | 6 | 200 | 1 |

**B. eSNP-eGene pairs among ct-eQTLs significant in both monocytes/ macrophages and microglia**
| eGene | eSNP | Position | MAF | Monocytes/Macrophages |  | Microglia |  | AD GWAS<br>P-value<sup>25</sup> |
| --- | --- | --- | --- | --- | --- | --- | --- | --- |
|  |  |  |  | Beta | P-value | Beta | P-value |  |
| HLA-DOB | rs3763355 | 6:32786882 | 0.06 | -2.02 | 9.98E-15 | 0.938 | 3.89E-14 | 0.001 |
| HLA-DOB | rs3763354 | 6:32786917 | 0.15 | -1.11 | 1.40E-10 | -0.642 | 2.80E-13 | 0.652 |
| HLA-DOB | rs1183595100 | 6:32768232 | NA | -1.13 | 8.34E-11 | -0.605 | 1.98E-11 | NA |

**C. Overlap of significant eQTLs in brain and blood with ct-eQTLs in myeloid cell types**
| Cell-types | Monocytes/ Macrophages |  | Microglia |  |
| --- | --- | --- | --- | --- |
|  | # ct-eQTLs<br>common to cell<br>type pair | # ct-eQTLs<br>unique to cell<br>type pair | # ct-eQTLs<br>common to cell<br>type pair | # ct-eQTLs<br>unique to cell<br>type pair |
| **Blood** |  |  |  |  |
| Neutrophils1 |  |  | 3 (0.3%)<sup>2</sup> | 0 |
| CD4+T-Cells |  |  | 3 (0.5%) | 0 |
| NK/CD8+T-Cells |  |  | 337 (12.9%) | 0 |
| Erythrocytes |  |  | 119 (5.6%) | 0 |
| Monocytes/ Macrophages |  |  | 251 (7.8%) | 3 |
| Unknown |  |  | 0 | 0 |
| Interferon/ Anti-bacterial |  |  | 628 (3.3%) | 63 |
| Neutrophils2 |  |  | 0 | 0 |
| B-cells |  |  | 0 | 0 |
| Eosinophils |  |  | 38 (5.2%) | 0 |
| **Brain** |  |  |  |  |
| Endothelial Cells | 55 ( 0.5%) | 0 |  |  |
| Neurons | 250 (1.3%) | 0 |  |  |
| Microglia | 251 (1.6%) | 3 |  |  |
| Astrocytes | 0 | 0 |  |  |
| Oligodendroglia | 0 | 0 |  |  |

<sup>2</sup> number in parentheses represent the proportion of ct-eQTLs for each cell type on the left that were also observed in either microglia or monocytes/macrophages

Considering significant eGene-eSNP pairs in myeloid cell types, 251 pairs including five distinct eGenes (*BTNL3, FAM118A, HLA-DOB, HLA-DRB1*, and *HLA-*DRB5) are shared between microglia and monocytes/macrophages (Table 4A and Fig. 2A). Three of these pairs involving eSNPs rs3763355, rs3763354, and rs1183595100 have the same target gene *HLA-DOB* and occur only in microglia and monocytes/macrophages (Table 4B).

**Figure 2.**
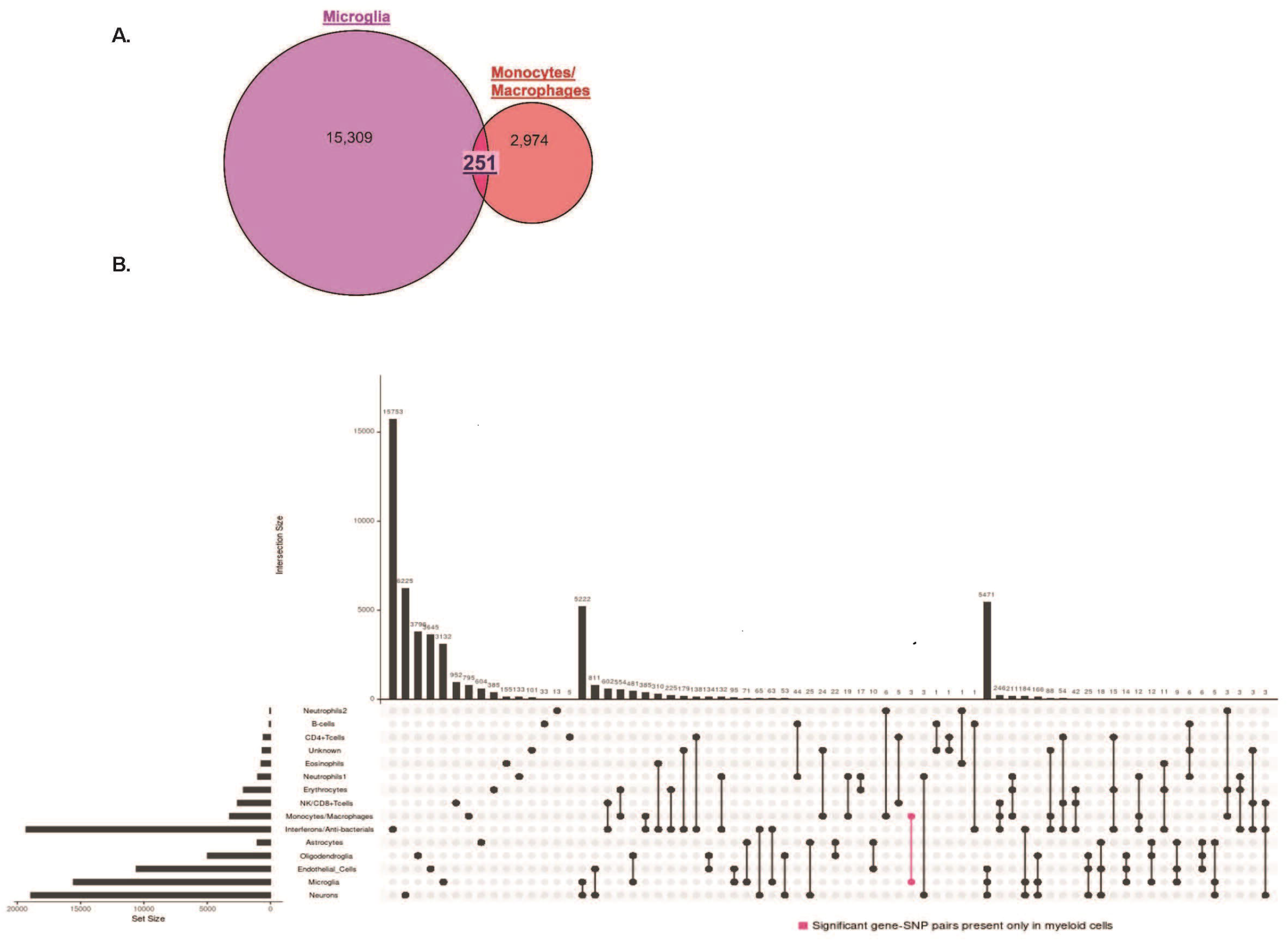
Intersection of significant gene-SNP eQTL pairs between cell-types in blood and brain tissue. **A)** Venn diagram showing overlap of ct-eQTL pairs in myeloid cell types (microglia and monocytes/macrophages). **B)** Number of significant eQTLs unique to and that overlap cell-types in blood and brain. Bar chart on the left side indicates the number of significant eQTLs involving each cell-type and bar chart above the matrix indicates the number of significant eQTLs that are unique to each cell type and set of cell-types. Pink colored bar indicates the number of eQTLs pairs that are unique to microglia and monocytes/macrophages.

Among the significant ct-eQTLs in brain, the cell types with the largest proportion that were also significant in monocytes/macrophages were microglia (1.6%) and neurons (1.3%) (Table 4C). Conversely, among the significant ct-eQTLs in blood, the cell types with the largest proportion that were also significant in microglia were NK/CD+ T-cells (12.9%) and monocytes/macrophages (7.8%). Among ct-eQTLs which are significant only for one cell-type each in blood and one in brain, monocytes/ macrophages shared three ct-eQTLs with microglia but with no other brain cell-types (Fig. 2B, Table 4C). By comparison, microglia shared 63 ct-eQTLs with interferons/ anti-bacterial cells, but with no other blood cell types. The much larger number of ct-eQTLs in microglia that were common with interferons/bacterial cells than monocytes/ macrophages may reflect the substantially greater proportion of significant eQTLs in blood involving interferons/ antibacterial cells (64%) than monocytes/macrophages (10.6%) (Table S10). The only other ct-eQTLs that were unique to a pair of cell types in brain and blood cell type involved neurons paired with neutrophils (n=3) and with interferons/anti-bacterial cells (n=65) (Fig. 2B).

## DISCUSSION

We identified several novel AD-related eQTLs that highlight the importance of cell-type dependent context. It is noteworthy that there were more significant ct-eQTLs in brain (n=51,098) than blood (n=30,405) even though the dataset containing expression data from blood (FHS) is several times larger than the brain expression dataset (ROSMAP). This could be due to greater cell type heterogeneity in brain, the enrichment of AD cases in the ROSMAP dataset who may show different patterns of gene expression compared to persons without AD, or highly variable gene expression across cell-types in the nervous system ^35^. Because expression studies in brain are often constrained by the small number specimens compared to studies in other tissues, post-mortem changes that may affect gene expression in brain ^36^, and the growing recognition that AD is a systemic disease ^37-39^, incorporating expression data from multiple tissues can enhance discovery of additional genetic influences on AD risk and pathogenesis.

Although most significant findings were tissue-specific, the 386 distinct eGenes among more than 24,000 significant gene-SNP eQTL pairs that were shared between blood and brain were enriched in the apoptosis signaling pathway that contributes to much of the underlying pathology associated with AD ^40, 41^. Five established AD genes (*CR1, ECHDC3, HLA-DRB1, HLA-DRB5*, and *WWOX* ^25, 28^) were shared eGenes in brain and blood and could be playing a key role in the systemic AD mechanisms. The complement receptor 1 (*CR1*) gene encodes a transmembrane glycoprotein functioning in the innate immune system by promoting phagocytosis of immune complexes, cellular debris, and Aβ ^42^. *CR1* is an eGene for several eSNPs, including AD GWAS peak SNP rs6656401 located within the gene, in brain and blood eQTLs and the effects on *CR1* expression are opposite in blood and brain. There are multiple possible explanations for the effect direction differences across tissues. The effect of eSNP rs6656401 on *CR1* expression may be developmental, noting that the average age of the FHS subjects (group with expression data in blood) is more than 30 years younger than the ROSMAP subjects (group with expression data in brain). The difference between brain and blood may also reflect post-mortem changes in brain that are not indicative of expression *in vivo*.

Alternatively, these effects may be related to AD because few FHS subjects were AD cases at the time of blood draw whereas 60% of subjects in the ROSMAP sample are AD cases. This idea is supported by the observation of a larger and positive effect of rs6656401 on *CR1* expression in AD (β=0.020) compared to control brains (β=-0.0086). Opposite effect directions of expression in brain and blood from AD patients compared to controls has been observed for several ribosomal genes ^43^. GWS variants located in the region spanning *ECHDC3* and *USP6NL* have previously been associated with AD ^44^. We found that *ECHDC3* is the target gene for eSNP rs866770710 located in its promoter region, and this eQTL was significant in brain and specifically in neurons. Altered *ECHDC3* expression in AD brains ^45^ supports the idea that this gene has a role in AD. Knockout of *WWOX* in mice leads to aggregation of amyloid-β (Aβ) and Tau, and subsequent cell death ^46, 47^.

The human leukocyte antigen (HLA) region is the key susceptibility locus in many immunological diseases and many associations have been reported between neurodegenerative diseases and HLA haplotypes ^48^. In addition, the most widely used marker to examine activated microglia in normal and diseased human brains is *HLA-DR* and microglia activation increases with the progression of AD ^49, 50^. *HLA-DRB5* and *HLA-DRB1* have been implicated in numerous GWAS studies as significantly associated with AD risk ^25, 28^ and appeared frequently among significant results in blood and brain in this study. Rs9271058, which is located approximately 17.8 kb upstream of *HLA-DRB1*, is significantly associated with AD risk (p=5.1 × 10^−8 25^) and when paired with *HLA-DRB1* is a significant eQTL and ct-eQTL in multiple cell types in blood and brain including myeloid lineage cells (i.e., monocytes/macrophages and microglia). This eSNP is also a significant eQTL in brain and specifically in neurons when paired with *HLA-DRB5*. Rs9271192, which is adjacent to rs9271058 and also significantly associated with AD risk (*P*=2.9 × 10^−12^) ^28^, is a significant eQTL and ct-QTL with multiple cell types in brain but not blood when paired with *HLA-DRB5 and HLA-DRB1*.

Significant associations for AD have been reported with variants spanning a large portion of the major histocompatibility (MHC) region in HLA class I, II and III loci ^48, 51, 52^. While the strongest statistical evidence for association in this region is with variants in *HLA-DRB1* ^25^, fine mapping in this region suggests that a class I haplotype (spanning the *HLA-A* and *HLA-B* loci) and a class II haplotype (including variants in *HLA-DRB1, HLA-DQA1* and *HLA-DQB1*) are more precise markers of AD risk. Given the complexity of the MHC region and extensive LD, further work is needed to confirm whether this is a true eQTL or a signal generated from a specific HLA allele or HLA haplotype. Although functional studies may be required to discern which HLA variants have AD-relevant consequences relevant to AD and develop methods to detect the differential expression of the HLA alleles, our findings support a role for the immune system in AD ^37, 53^ and the hypothesis that a large proportion of AD risk can be explained by genes expressed in myeloid cells ^10^.

The potential importance and relevance to AD of eQTLs and ct-eQTLs in myeloid cell-types is supported by the observation that a large portion of GWS ct-eQTLs we identified map within 1 Mb of established AD loci, and 58% (12/20 in monocytes/ macrophages and 11/20 in microglia) of the most significant eGenes have been previously implicated in AD. *DLG2* encodes a synaptic protein whose expression was previously reported as down-regulated in an AD proteome and transcriptome network ^54^ and inversely associated with AD Braak stage ^30^. Genome-wide significant associations of AD risk with *PTPRG* was observed in a family-based GWAS ^55^ and with *CLNK* in a recent large GWAS for which the evidence was derived almost entirely with a proxy AD phenotype in the UK Biobank ^56^. *NFXL1* is a novel putative substrate for *BACE1*, an important AD therapeutic target ^57^. *FCRL5* may interact with the *APOE*E2* allele and also modifies AD age of onset ^58^. *C4BPA* was shown to be a consistently down-regulated in MCI and AD patients, and the protein encoded by this gene accumulates in Aβ plaques in AD brains ^31, 59^. Lower levels of the *PAM* have been observed in the brains and CSF of AD patients compared to healthy controls ^60^ and *MYO1E* is expressed by anti-inflammatory disease associated microglia ^32^. As a calcium channel protein, *CACNB2* may affect AD risk by altering calcium levels which could cause mitochondrial damage and then induce apoptosis ^61, 62^.

Likewise, several eGenes of top-ranked ct-eQTLs in microglia that are not established AD loci may have a role in the disease. It was shown that copy number variants (CNVs) near *HNRNPCL1* overlapped the coding portion of the gene in AD cases but not controls ^63^. A region of epigenetic variation in *ALLC* was associated with AD neuropathology ^33^. *FAM21B*, a retromer gene in the endosome-to-Golgi retrieval pathway, was associated with AD in a candidate gene study ^64^. Vacuolar sorting proteins genes in this pathway including *SORL1* have been functionally linked to AD through trafficking of Aβ ^65^. One study demonstrated that *WNT3* expression in the hippocampus was increased by exercise and alleviated AD-associated memory loss by increasing neurogenesis ^34^. Expression of *RPL9* is downregulated in severe AD ^66^ and significantly differs by sex among persons with the *APOE* □4 allele ^67^. Significant evidence of association with a *TRIM49B* SNP was found in a genome-wide pleiotropy GWAS of AD and major depressive disorder (MDD) ^68^.

*HLA-DOB*, which is one of the five distinct eGenes (*BTNL3, FAM118A, HLA-DOB, HLA-DRB1*, and *HLA-DRB5)* for significant ct-eQTLs shared between microglia and monocytes/macrophages, and is the target gene for three eSNPs (rs3763355, rs3763354, and rs1183595100) that were evident only in these myeloid cell types.

These eSNPs have similar eQTL p-values in both cell types, but have slightly larger effect sizes in monocytes (Fig. 2). The effect of rs3763355 on expression is in opposite directions in monocytes and microglia which suggests *HLA-DOB* may be acting in different immune capacitates in AD in blood and brain. Though the functions of the genes *BTNL3* and *FAM118A* are unknown, a *BTNL8-BTNL3* deletion has been correlated with TNF and ERK1/AKT pathways, which have an important role in immune regulation inducing inflammation, apoptosis, and proliferation, suggesting the deletion could be correlated to inflammatory disease ^69^. This suggests that the majority of the shared myeloid cell types genes-the *HLA* genes and possibly *BTNL3*, are all immune-related. Ct-eQTLs involving microglia and monocytes/macrophages had a larger proportion of total intersection, an isolated set interaction and a statistically significant overlap (*P*<1.0E-314), demonstrating a stronger connection than other brain/blood cell types in this study and thus providing further evidence for importance of the immune system in AD.

The proportions of significant ct-eQTLs in NK cells/CD8+T cells, monocytes/ macrophages, and eosinophils are comparable to those observed in reference blood tissue ^70, 71^. Similarly, significant eQTL distributions in endothelial cells, neurons, and glia are consistent with reference brain tissue ^72^. The majority of significant blood eQTLs were type I interferon response cells which cross-regulate with pro-inflammatory cytokines that drive pathogenesis of autoimmune diseases including AD and certain heart diseases ^73-75^and the enrichment of interferon ct-eQTLs in this study could possibly be due to the high proportion of subjects these diseases in the FHS dataset. In contrast, the proportion of significant ct-eQTLs among glial cells is much lower in astrocytes and oligodendrocytes and three-fold higher in microglia than in reference brain tissue ^72^. Because many AD risk genes are expressed in myeloid cells including microglia ^10^, the large number of microglia ct-eQTLs is consistent with the high proportion of AD subjects in the ROSMAP dataset.

Several SNPs previously reported to be associated with AD at the GWS level were associated with eGenes that differ from genes ascribed to AD loci and thus may have a role in AD. Karch et al. observed that the expression of *PILRB*, which is involved in immune response and is the activator receptor to its inhibitory counterpart *PILRA*, an established AD gene ^76, 77^, was highest in microglia ^11^. *CNN2*, the eGene for eSNP rs4147929 located near the end of *ABCA7*, significantly alters extracellular Aβ levels in human induced pluripotent stem cell-derived neurons and astrocytes ^78^. Rs4147929 also targeted *HMHA1* which plays several roles in the immune system in an HLA-dependent manner ^79^. The eSNP/GWAS SNP rs3740688 located in *SPI1* also affects expression of *MYBPC3* and *MADD. MYBPC3* was recently identified as a target gene for eSNPs located in *CELF1* and *MS64A6A* in a study of eQTLs in blood for GWS AD loci ^80^. *MADD* is expressed in neurons ^11^, is involved in neuronal cell death in the hippocampus ^81^, and was shown to be a tau toxicity modulator ^82^. Although eSNP rs113986870 in *KANSL1* when paired with the nearby eGene *LRRC37A2* was a significant brain eQTL and ct-eQTL, *LRRC37A2* encodes a leucine rich repeat protein that is expressed primarily in testis and has no apparent connection to AD. However, rs113986870 also significantly influenced expression of another gene in this region, *ARL17A*, that was previously linked to progressive supranuclear palsy by analysis of gene expression and methylation ^83^.

Our study has several noteworthy limitations. The proxy genes individually or collectively may not accurately depict cell-type specific context. In addition, the comparisons of gene expression in blood and brain may yield false results because they are based on independent groups ascertained from a community-based longitudinal study of health (FHS – blood) and multiple sources for studies of AD (ROSMAP – brain) which were not matched for age, sex, ethnicity and other factors which may affect gene expression. Moreover, the FHS cohort contains many elderly but relatively few AD cases, whereas nearly 60% of the ROSMAP participants in this autopsy sample are AD cases. Although the dataset for eQTL analysis in blood was much larger than the dataset derived from brain, the effect sizes associated with many of the eQTLs common to both tissues were similar. Also, findings in brain may reflect post-mortem changes unrelated to disease or cell-type different expression ^36^. Another limitation of our findings is that some cell types are vastly under-represented compared to others.

Because myeloid cell types are represented in only a small proportion of the total cell populations in brain and blood (generally ∼10%), it is difficult to identify myeloid-specific signals ^12^. Despite this limitation, many of the most significant and noteworthy results of this study involved monocytes/macrophages and microglia. Finally, some of the findings may not be AD-related because expression was not compared between AD cases and controls.

## Conclusion

Our observation of cell-type specific expression patterns for established and potentially novel AD genes, finding of additional evidence for the role of myeloid cells in AD risk, and discovery of potential novel blood and brain AD biomarkers highlight the importance of cell-type specific analysis. Future studies that compare cell type specific differential gene expression among AD cases and controls using single cell RNA-sequencing or cell count data, as well as functional experiments, are needed to validate and extend our findings.

## Supporting information

Supplemental resources

## Data Availability

This research was conducted in part using data and resources from the FHS of the National Heart, Lung, and Blood Institute (NHLBI) of the US National Institutes of Health (NIH) and the Boston University School of Medicine. The analyses reflect intellectual input and resource development from the FHS investigators participating in the SNP Health Association Resource (SHARe) project and in the Systems Approach to Biomarker Research in Cardiovascular Disease (SABRe) project.
The results published here are in whole or in part based on data obtained from the AD Knowledge Portal (https://adknowledgeportal.synapse.org). Study data were provided by the Rush Alzheimers Disease Center, Rush University Medical Center, Chicago. Data collection was supported through funding by NIA grants P30AG10161 (ROS), R01AG15819 (ROSMAP; genomics and RNAseq), R01AG17917 (MAP), R01AG30146, R01AG36042 (5hC methylation, ATACseq), RC2AG036547 (H3K9Ac), R01AG36836 (RNAseq), R01AG48015 (monocyte RNAseq) RF1AG57473 (single nucleus RNAseq), U01AG32984 (genomic and whole exome sequencing), U01AG46152 (ROSMAP AMP-AD, targeted proteomics), U01AG46161(TMT proteomics), U01AG61356 (whole genome sequencing, targeted proteomics, ROSMAP AMP-AD), the Illinois Department of Public Health (ROSMAP), and the Translational Genomics Research Institute (genomic). Additional phenotypic data can be requested at www.radc.rush.edu.

## ACKNOWLEDGEMENTS

This study was supported by NIH grants RF1-AG057519, 2R01-AG048927 U01-AG058654, P30-AG13846, 3U01-AG032984, U01-AG062602 and U19-AG068753. Framingham brain bank data was supported by grants 75N92019D00031 and HHSN2682015000011. Collection of study data provided by the Rush Alzheimer’s Disease Center, Rush University Medical Center, Chicago was supported through funding by NIA grants P30AG10161, R01AG15819, R01AG17917, R01AG30146, R01AG36836, U01AG32984, U01AG46152, U01AG61358, a grant from the Illinois Department of Public Health, and the Translational Genomics Research Institute.

