## Supplemental materials for "Cell-type Specific Expression Quantitative Trait Loci Associated with Alzheimer Disease in Blood and Brain Tissue"

**Table S1.** Characteristics of subjects in the FHS and ROSMAP datasets

**Table S2.** Proxy genes for cell types in blood and brain

**Table S3.** Genome-wide summary of eQTLs and ct-eQTLs in blood and brain

**Table S4.** Established AD loci

**Table S5.** Target genes of significant eQTLs and ct-eQTLs shared between blood and brain

**Table S6.** Pathway analysis of eGenes shared between blood and brain

**Table S7.** eGenes shared by eQTLs and ct-eQTLs in blood and brain

**Table S8.** Established AD genes among significant eQTLs and ct-eQTLs

**Table S9.** Significant eGenes located in AD association peaks

**Table S10.** Cell-type distribution of significant ct-eQTLs

**Table S11.** Gene-set enrichment analysis including eGenes of significant ct-eQTLs in myeloid cell lineages

**Fig. S1.** Study design

**Fig. S2.** Regional plots for colocalized AD GWAS/lead eQTL variant pairs

**Supplemental Resources.** Top-ranked eSNPs per eGene for significant eQTLs and ct-eQTLs in blood and brain.

**Table S1:** Characteristics of subjects in the Framingham Heart Study (FHS) and Religious Orders Study (ROS)/ Memory and Aging Project (MAP) datasets

| **Dataset** | **Race** | **N** | **AD Cases** | **Controls** | **Males** | **Females** | **Mean Age (SD) ^1^** | **% *APOE* ε4 carrier** |
| --- | --- | --- | --- | --- | --- | --- | --- | --- |
| FHS | European Ancestry | 5,257 | 104(2%) | 5,153 (98%) | 2,421(46%) | 2,836 (54%) | 54.9 (13.3) | 23% |
| ROSMAP | 98% Caucasian,  2% African American,  < 0.01 % other | 475 | 281(59%) | 194 (41%) | 175 (37%) | 300 (63%) | 85.9 (4.8) | 26% |

^1^Age at onset for AD cases, age at exam for controls

**Table S2**: Proxy genes for cell types in blood and brain

| **Proxy gene** | **Tissue** | **Cell-type** | **Function/GO biological process** |
| --- | --- | --- | --- |
| STX3 | Blood | Neutrophils 1 | Detection of bacterium |
| FAM102A | Blood | CD4+ T-Cells | T cell selection |
| SAMD3 | Blood | NK cells / CD8+ T-Cells | Cellular defense response |
| TSPAN5 | Blood | Erythrocytes | Hemoglobin metabolic process |
| PATL1 | Blood | Monocytes / Macrophages | Defense response to virus |
| PACS1 | Blood | Unknown | Unknown/ Nerve growth factor receptor signaling pathway |
| SP140 | Blood | Interferon response(+)/ Anti-bacterial(-) | Type 1 interferon response/Anti-bacterial, Regulation of defense response |
| FBXL5 | Blood | Neutrophils 2 | Detection of bacterium |
| STRBP | Blood | B-cells | B cell receptor signaling pathway |
| SIGLEC8 | Blood | Eosinophils | Regulation of myeloid leukocyte mediated immunity |
| CD34 | Brain | Endothelial Cells | Lining blood vessels |
| RELN | Brain | Neurons | Main signaling units |
| CD68 | Brain | Microglia | Brain immune cells |
| GFAP | Brain | Astrocytes | Support neuronal growth, function and neurotransmitter recycling |
| OLIG2 | Brain | Oligodendroglia | Insulating neuronal axons |

**Table S3:** Genome-wide summary of cis eQTLs and ct-eQTLs in blood and brain

|  | **Blood eQTL** | **Blood ct-eQTL** | **Brain eQTL** | **Brain ct-eQTL** |
| --- | --- | --- | --- | --- |
| Total # of tests | 8,662,143 | 86,621,430 | 304,732,096 | 1,523,660,480 |
| # significant eQTL pairs | 847,429 | 30,405 | 173,857 | 51,098 |
| p-value threshold ^1^ | 5.77E-09 | 5.77E-10 | 1.64E-10 | 3.28E-11 |
| # unique eGenes in eQTL pairs | 6033 | 502 | 1,301 | 799 |
| # unique eSNPs in eQTL pairs | 727,384 | 22,226 | 144,258 | 26,757 |

**^1^** Bonferroni corrected

**Table S4:** Established AD loci

| **Chr** | **Pos (hg37)** | **rsID** | **Nearest Gene** | **Method^1^** | **Trait^2^** | **p-value** | **Ref** |
| --- | --- | --- | --- | --- | --- | --- | --- |
| 1 | 1:207692049 | rs6656401 | CR1 | G | AD | 5.7 × 10^−24^ | [1] |
| 1 | 1:227058273 | N/A | PSEN2 | L,C | AD | NA | [2, 3] |
| 2 | 2:106642554 | rs34487851 | ECRG4 | G | NP + NFT | 2.4 × 10^−8^ | [4] |
| 2 | 2:127892810 | rs6733839 | BIN1 | G | AD | 6.9 × 10^−44^ | [1] |
| 2 | 2:202149628 | rs146286958 | CASP8 | RVG | AD | 8.6 x 10^-5 a^ | [5] |
| 2 | 2:234068476 | rs35349669 | INPP5D | G | AD | 3.2 × 10^−8^ | [1] |
| 2 | 2:3474085 | rs35067331 | TRAPPC12 | G | NFT + CAA | 5.8 × 10^−8^ | [4] |
| 2 | NA | NA | ADI1 | GB | NFT + CAA | <1 × 10^−6^ ^b^ | [4] |
| 3 | 3:178257562 | rs9637454 | KCNMB2 | G | AD | 7.1 × 10^−8^ | [6] |
| 3 | 3:190951729 | rs9877502 | OSTN | G | CSF tau | 4.9 × 10^−9^ | [7] |
| 4 | 4:174094940 | rs62341097 | GALNT7 | G | NP | 6.0 × 10^-9^ | [6] |
| 4 | 4:7353052 | rs13110208 | SORCS2 | CG,F | AD | 9.0 × 10^-3^ | [8] |
| 4 | 4:95170280 | rs137875858 | UNC5C | L,CG,F | AD | 9.5 × 10^-3 c^ | [9] |
| 5 | 5:139707439 | rs11168036 | PFDN1,  HBEGF | G | AD | 7.1 × 10^−9^ | [10] |
| 5 | 5:15669858 | rs75002042 | FBXL7 | G | AD | 6.2 x 10^-9^ | [11] |
| 5 | 5:88223420 | rs190982 | MEF2C | G | AD | 3.2 × 10^−8^ | [1] |
| 6 | 6: 83072923 | rs547721664 | TPBG | G | AD | 1.8 × 10^−6^ | [10] |
| 6 | 6:32575406 | rs9271058 | HLA-DRB1 | G | AD | 5.1 × 10^−8^ | [12] |
| 6 | 6:32578530 | rs9271192 | HLA-DRB5 | G | AD | 2.9 × 10^−12^ | [1] |
| 6 | 6: 41034000 | rs114812713 | OARD1 | G | AD | 2.1x10^-13^ | [6] |
| 6 | 6:41129252 | rs75932628 (R47H) | TREM2 | G,F | AD | 2.9 × 10^−12^ | [7, 12] |
| 6 | 6:41186912 | rs9381040 | TREML2 | G,F | AD | 6.3 × 10^−7^ | [1, 7] |
| 6 | 6:41368363 | rs6922617 | NCR2 | G | AD | 3.6 × 10^−8^ | [7] |
| 6 | 6:47487762 | rs10948363 | CD2AP | G | AD | 5.2 × 10^−11^ | [1] |
| 7 | 7:100004446 | rs1476679 | ZCWPW1 | G | AD | 5.6 × 10^−10^ | [1] |
| 7 | 7:132110923 | rs277470 | PLXNA4 | G | AD | 4.1 × 10^-8^ | [13] |
| 7 | 7:143110762 | rs11771145 | EPHA1 | G | AD | 1.1 × 10^−13^ | [1] |
| 7 | 7:154988675 | NA | AC099552.4 | RVG | AD | 1.2 × 10^−7 d^ | [14] |
| 7 | 7:18698331 | rs79524815 | HDAC9 | G | NFT + CAA | 1.1 × 10^−8^ | [4] |
| 7 | 7:37841534 | rs2718058 | NME8 | G | AD | 4.8 × 10^−9^ | [1] |
| 7 | 7:51578022 | rs112404845 | COBL | G | AD | 3.8 × 10^-8^ | [15] |
| 7 | 7:88406552 | rs73705514 | ZNF804B | G | LMdT | 2.9 × 10^−9^ | [16] |
| 7 | 7:91709085 | rs144662445 | AKAP9 | RVG,F | AD | 2.2 x 10^-3 c^ | [17] |
| 7 | NA | NA | PILRA | GB | AD | 2.3 × 10^−6 e^ | [18, 19] |
| 8 | 8:145138063 | rs138412600 | GPAA1 | G | AD | 7.8 × 10^-8 f^ | [20] |
| 8 | 8:17496561 | rs4921790 | PDGFRL / MTUS1 | G | HPV | 4.6 × 10^−9^ | [16] |
| 8 | 8:27195121 | rs28834970 | PTK2B | G | AD | 7.4 × 10^−14^ | [1] |
| 8 | 8:27467686 | rs9331896 | CLU | G | AD | 2.8 × 10^−25^ | [1] |
| 8 | 8:95958637 | rs1713669 | TP53INP1 | GB | AD | 1.4 × 10^−6 g^ | [21] |
| 9 | 9: 3929424 | rs514716 | GLIS3 | G | CSF tau | 3.2 ×10^−9^ | [7] |
| 10 | 10:107013252 | rs7920533 | SORCS3 | CG,F | AD | 1.0 × 10^-2^ | [8] |
| 10 | 10:108562008 | rs12248379 | SORCS1 | CG,F | AD | 3.0 × 10^-3^ | [8] |
| 10 | 10:115489177 | rs1116437863 | CASP7 | G | AD | 2.4 × 10^-10^ | [22] |
| 10 | 10:11720308 | rs7920721 | ECHDC3 | G | AD | 2.3 × 10^−9^ | [12] |
| 10 | 10:11720308 | rs7920721 | USP6NL | G | AD | 3.0 × 10^−8^ | [10] |
| 10 | 10:13966445 | rs2446581 | FRMD4A | G | AD | 1.1 × 10^−10^ | [23] |
| 11 | 11:121435587 | rs11218343 | SORL1 | G,CG | AD | 2.7 × 10^−8^ | [1, 12, 24] |
| 11 | 11:47380340 | rs3740688 | SPI1 | G | AD | 9.7 × 10^−11^ | [12] |
| 11 | 11:47557871 | rs10838725 | CELF1 | G | AD | 1.1 × 10^−8^ | [1] |
| 11 | 11:59923508 | rs983392 | MS4A6A | G | AD | 6.1 × 10^−16^ | [1] |
| 11 | 11:59936926 | rs7933202 | MS4A2 | G | AD | 2.2 × 10^−15^ | [12] |
| 11 | 11:60021948 | rs1582763 | MS4A4A | G | AD | 1.15 × 10^-15^ | [25] |
| 11 | 11:85867875 | rs10792832 | PICALM | G | AD | 9.3 × 10^−26^ | [1] |
| 11 | NA | NA | OR8G5 | GB | AD | 4.7 × 10^−7 h^ | [20] |
| 12 | 12:119390525 | rs10775009 | SRRM4 | G | CSF Tau | 1.6 × 10^−9^ | [16] |
| 13 | 13:103663945 | rs16961023 | SLC10A2 | G | AD | 4.6 × 10^-8^ | [15] |
| 14 | 14:105385352 | rs2819438 | PLD4 | G | CSF Tau | 6.9 × 10^−9^ | [16] |
| 14 | 14:106236128 | rs12890621 | IGHG3 | G | AD | 9.8 × 10^−7^ | [14] |
| 14 | 14:106680831 | rs2011167 | IGHV1-67 | G | AD | 7.9×10^−8 g^ | [21] |
| 14 | 14:53400629 | rs17125944 | FERMT2 | G | AD | 7.9 × 10^−9^ | [1] |
| 14 | 14:73637653 | rs63749824 | PSEN1 | L,C | AD | NA | [26] |
| 14 | 14:92926952 | rs10498633 | SLC24A4 / RIN3 | G | AD | 5.5 × 10^−9^ | [1] |
| 14 | 14:92932828 | rs12881735 | SLC24A4 | G | AD | 7.4 × 10^−9^ | [12] |
| 14 | NA | NA | IGHV3-7 | GB | AD | 9.75 × 10^−16^ | [20] |
| 15 | 15:101646763 | rs139709573 | TM2D3 | RVG | AD | 6.6 × 10^-9^ | [27] |
| 15 | 15:59045774 | rs593742 | ADAM10 | G | AD | 6.8 × 10^−9^ | [12] |
| 15 | 15:64433291 | rs74615166 | TRIP4 | G | AD | 9.7 × 10^−9^ | [28] |
| 16 | 16:19808163 | rs7185636 | IQCK | G | AD | 2.4 × 10^−8^ | [12] |
| 16 | 16:79355857 | rs62039712 | WWOX | G | AD | 3.7 × 10^−8^ | [12] |
| 16 | 16:81942028 | rs72824905 | PLCG2 | G | AD | 5.4 × 10^−10^ | [29] |
| 17 | 17:44353222 | rs2732703 | MAPT | G | AD | 5.8 × 10^−9^ | [30] |
| 17 | 17:44355683 | rs113986870 | KANSL1 | G | AD | 1.3 × 10^−8^ | [30] |
| 17 | 17:47297297 | rs616338 | ABI3 | G | AD | 4.6 × 10^−10^ | [29] |
| 17 | 17:56409089 | rs2632516 | BZRAP1 | G | AD | 4.4×10^−8^ | [10] |
| 17 | 17:61569732 | rs4351 | ACE | G | AD | 5.3×10^-9^ | [31] |
| 19 | 19:1063443 | rs4147929 | ABCA7 | G | AD | 1.1 × 10^−15^ | [1] |
| 19 | 19:15302421 | rs149307620 | NOTCH3 | RVG,F | AD | NA | [32] |
| 19 | 19:18546678 | rs2303697 | ISYNA1 | G | AD | 4.6 × 10^−7 f^ | [20] |
| 19 | 19:40877595 | rs145999145 | PLD3 | RVG,F | AD | 1.4 × 10^-11^ | [33] |
| 19 | 19:45411941 | rs429358 | APOE | L,CG | AD | 2.1 × 10^−47^ | [34] |
| 19 | 19:51727962 | rs3865444 | CD33 | G | AD | 1.6 × 10^−9^ | [35] |
| 20 | 20:55018260 | rs7274581 | CASS4 | G | AD | 2.5 × 10^−8^ | [1] |
| 20 | NA | NA | SLC24A3 | GB | AD | 2.7 × 10^−12^ | [20] |
| 21 | 21:28915457 | rs1487586185 | APP | CG | AD | NA | [36] |
| 21 | 21:28156856 | rs2830500 | ADAMTS1 | G | AD | 2.6 × 10^−8^ | [12] |
| 21 | 21:43678066 | rs142544282 | ABCG1 | G | NP | 8.0×10^−9^ | [6] |

^1^**Method:** G = GWAS; RVG = Rare Variant GWAS; C =Cloning; CG = Candidate Gene; L = Linkage; F =Functional evidence; GB = Gene-based;

^2^**Trait:** AD = Alzheimer disease, HPV = hippocampal volume, LMdT = logical memory – delayed recall, NFT = neurofibrillary tangle, NP = neuritic plaque

Study-wide significance level: ^a^ P=8x10^-4^, ^b^ P=2.7x10^-6^; ^c^ P=0.05, ^d^ P=3.1x10^-7^, ^e^ P=0.0014, ^f^ P=2.8x10^-7^, ^g^ P=2.5x10^-6^, ^h^ P=6.4x10^-7^,

**Table S5:** Target genes of significant eQTLs and ct-eQTLs shared between blood and brain

| ACACA  ACCS  ACOT4  ADAL  ADAT1  ADHFE1 ADORA2B AGA  AHI1  AIFM2  AK5  ALDH8A1  AMACR ANAPC4 ARHGEF3 ARHGEF35 ARL17A AS3MT ASRGL1  ATF6  ATP5S ATP6V1E2 ATXN7L3B B3GNTL1 BATF3  BLMH  BLOC1S2 BLVRA  BNIP1  BTN2A2 BTNL3  C12orf60 C14orf166 C15orf57 C1QTNF6 C21orf128 C22orf34 C7orf13  C8orf31 | C9orf72 C9orf89 CAB39L  CAT  CBLN3  CBR3  CCBL2  CCDC13 CCDC170  CCDC23 CCDC25 CCDC82 CD274  CD6  CDA  CDC25B  CDC7  CDK10 CENPP CENPV CEP19 CERS5 CHAF1A CHCHD2 CHKB-CPT1B CHRNA5 CHRNE  CKS2  CLTCL1 CNDP2 CNGA1  CPVL  **CR1 ***  CRIPT  CRLF3  CRYZ CSGALNACT1  CSGALNACT2  CTNS | CYSTM1 DCAKD DCBLD2  DDT  DHRS1 DNAJC15 DPM2  DSC1  DSCC1 DUSP14 **ECHDC3 *** EFCAB13 EFCAB2 EFHB  EIF2A ENDOG  EPG5  ERAP2  ERCC3  ERMAP ETFDH  ETS2  ETV7 EXOSC6 EYA3  F2RL1  F5  FADD  FAH FAM118A FAM3B FANK1  FAS  FBN2  FEM1A  FEZ2  FIGNL1  FLT3 FLYWCH1 | FOPNL  FOXRED1 FOXRED2 FRA10AC1 FUT2  FXN  GAA  GALC  GATC  GBP3  GCLM GGNBP2 GIMAP5 GLIPR1L2 GNL3  GNLY GOSR1  GP6  GPNMB GSR  GSTA1 GSTM3 GSTT1 GTSF1 GUF1 HAUS4 HBS1L HEATR3  HIATL1  HIBCH HIST1H3E HIST1H4C HIST1H4F HLA-DOB HLA-DQA2 HLA-DQB2 **HLA-DRB1* HLA-DRB5***  HP | HPR HSD17B12 HSD17B13 HTATIP2 HYAL3  IFI27  IFI27L1 IFI27L2 IFIT5  IFT46 IGFLR1 IL10RB IL18R1 IL1RL1  IL32  INPP1  IPMK  IQCB1  IQCG IQGAP1 ISCU  ITGB2  ITIH4 KANSL2 KIAA1430 KLC3 KLHDC4 KLKB1 KRR1 L3HYPDH L3MBTL3 LACTB  LDHC  LIG3  LILRB1 LIMD1  LNPEP LONP1  LPIN1 | LPIN2 LRPAP1 LRRC2 LRRC27 LRRC61 LRRCC1 LRRIQ3 LSG1  LXN  MAEL MAN2B2 MANBA MCFD2 MCM8 MCOLN2 MCPH1 MEI1 METTL18 METTL21B MGMT  MGRN1  MICB  MLH3 MON1B MPHOSPH6 MPPE1 MRPL10 MRPL18 MRPL19 MRPL21 MRPL53 MRPS10 MSH3  MSH4 MTHFS MTRR  MUL1  MZT<2A  NAPRT1 | NARS2 NDUFAF1 NIPSNAP3A NMRK1 NMUR1 NOP10 NPHP3  NRBF2 NSUN2 NSUN6 NUP107 NUP210L NUPL2 NUSAP1 OSCP1 PAAF1 PADI4  PAX8 PCNXL4 PDCD1LG2 PDHB PDLIM5 PDZK1IP1 PEX6  PIGN  PISD PLEKHH2 POLR1B POLR1E POLR2J POP5  PPA2 PPFIA1 PPIL3 PPM1N PPP2R1B  PTGR1 QRSL1 RABEP1 | RAD51C RARRES1 RBL2 RBPMS2 RCBTB1 RFWD3 RMI2 RNF166 RNPEP RPA2 RPL36AL RPP21 RRP1B RTN4  RWDD2B RWDD3  SAAL1 SCIMP SCLY  SELL  SETD4 SHISA4 SLC18A1 SLC22A18 SLC25A1 SLC25A24 SLC25A51 SLC26A8 SLC35A1  SLFN5 SMARCB1 SMC1B SMC2 SNX19 SNX32  SOHLH2 SPAG7 SPATA5L1 SPATA7 | SPECC1 SPEF2 SPPL3 SPSB2 SSR1 STYXL1 SULT1A1 SUMF1 SUPT3H SUPT4H1 SUSD1 TAF1C TAS2R4 TBC1D9B TC2N TCEA3 TCF19 TDRD6 TECPR1 TEN1 TESK2 TFB1M  THNSL2 TIMM10 TMEM156  TMEM245 TMOD3 TNFRSF10C TNFRSF13C TNNI3 TOMM7 TOP3B TRAPPC4  TREML4  TRIM35  TRIM58 TTC21B TVP23C UBALD2 | UFSP2 UHRF1BP1 ULK4 VN1R1 WARS2 WBP2NL WBSCR27 WDR27 WDR52 WDYHV1 WFDC3 WIPI1 **WWOX *** XRCC6BP1 XRRA1 YEATS4 YWHAB ZADH2 ZFP82 ZMAT3 ZMYND12 ZNF155 ZNF354C ZNF467 ZNF471 ZNF501 ZNF502 ZNF514  ZNF577 ZNF593  ZNF670, ZNF839 ZNRD1  ZP3  ZXDC |
| --- | --- | --- | --- | --- | --- | --- | --- | --- | --- |

**Table S6:** Pathway analysis of eGenes shared between blood and brain

| **PANTHER Pathway** | **# Genes**  **Annotated**  **to Pathway** | **# Genes**  **in Network** | **Expected**  **P-value *** | **Fold**  **Enrichment** | **Direction** | **Unadjusted**  **P-value** |
| --- | --- | --- | --- | --- | --- | --- |
| Apoptosis signaling pathway | 115 | 6 | 2.12 | 2.83 | + | 2.28E-02 |
| Wnt signaling pathway | 317 | 1 | 5.84 | 0.17 | - | 3.36E-02 |
| General transcription by RNA polymerase I | 17 | 2 | 0.31 | 6.39 | + | 4.55E-02 |

PANTHER = Protein ANalysis THrough Evolutionary Relationships) Classification System

*Expected probability of observing at least x number of genes out of the total n genes in the PANTHER list annotated to a particular pathway, given the proportion of genes in the reference Homo Sapiens whole genome that are annotated to that pathway

**Table S7:** eGenes shared by eQTLs and ct-eQTLs in blood and brain

| AS3MT  ATXN7L3B  BTNL3  DNAJC15 | EFCAB2  ERAP2  FAM118A  GSTT1 | HLA-DOB  **HLA-DRB1 ***  **HLA-DRB5 ***  HP | MRPL21  NMRK1  TIMM10  XRRA1 |
| --- | --- | --- | --- |

* Previously associated with AD risk by GWAS

**Table S8**: Established AD genes among significant eQTLs and ct-eQTLs

| **eGene** | **Tissue** | **Cell-type** | **Lead eSNP** | | **Position** | | **MAF** | | **Beta** | | **Std Error** | | **P-value** | | **# of total significant eSNPs in gene/cell-type** | | **Distance (bp) between eSNP and eGene**  **[Nearest gene]** |
| --- | --- | --- | --- | --- | --- | --- | --- | --- | --- | --- | --- | --- | --- | --- | --- | --- | --- |
| CR1 | Blood | NA | rs7533408 | | 1:207673631 | | 0.25 | | 0.059 | | 0.006 | | 3.60E-22 | | 169 | | 0 |
| PSEN2 | Blood | NA | rs1289395 | | 1:227042462 | | 0.37 | | -0.020 | | 0.003 | | 2.77E-12 | | 49 | | 15423 |
| TRAPPC12 | Blood | NA | rs71281795 | | 2:3487331 | | NAV | | -0.040 | | 0.005 | | 1.10E-15 | | 104 | | 0 |
| ADI1 | Blood | NA | rs57139325 | | 2:3519283 | | 0.18 | | 0.127 | | 0.008 | | 8.45E-58 | | 123 | | 0 |
| BIN1 | Blood | NA | rs1060743 | | 2:127826533 | | 0.29 | | -0.060 | | 0.003 | | 2.48E-99 | | 355 | | 0 |
| CASP8 | Blood | NA | rs7560328 | | 2:202164837 | | 0.39 | | 0.028 | | 0.003 | | 1.06E-18 | | 54 | | 12403 [In FLACC1] |
| INPP5D | Blood | NA | rs7581787 | | 2:234077240 | | 0.44 | | 0.026 | | 0.003 | | 5.79E-16 | | 158 | | 0 |
| GALNT7 | Blood | NA | rs1006003 | | 4:174092791 | | 0.50 | | -0.034 | | 0.003 | | 5.07E-24 | | 102 | | 0 |
| HLA-DRB5 | Blood | NA | rs9269008 | | 6:32436217 | | 0.17 | | -2.580 | | 0.057 | | <1.0E-314 | | 72 | | 48903 [HLA-DRB9 (5060)] |
| HLA-DRB1 | Blood | NA | rs9270815 | | 6:32569859 | | 0.14 | | -5.430 | | 0.044 | | <1.0E-314 | | 630 | | 12234 |
| TREML2 | Blood | NA | rs6933231 | | 6:41163700 | | 0.37 | | -0.034 | | 0.004 | | 5.39E-16 | | 34 | | 0 |
| CD2AP | Blood | NA | rs4711880 | | 6:47480676 | | 0.25 | | -0.146 | | 0.007 | | 1.36E-104 | | 331 | | 0 |
| NME8 | Blood | NA | rs71527594 | | 7:37875986 | | NAV | | 0.117 | | 0.006 | | 1.84E-81 | | 304 | | 12213 |
| EPHA1 | Blood | NA | rs3935067 | | 7:143104331 | | 0.33 | | 0.039 | | 0.003 | | 5.29E-34 | | 50 | | 0 |
| MTUS1 | Blood | NA | rs117496663 | | 8:17660151 | | 0.02 | | -0.290 | | 0.009 | | 9.94E-218 | | 325 | | 1725 |
| PTK2B | Blood | NA | rs28834970 | | 8:27195121 | | 0.34 | | -0.068 | | 0.003 | | 1.48E-100 | | 339 | | 0 |
| CLU | Blood | NA | rs9331950 | | 8:27454682 | | 0.22 | | -0.059 | | 0.010 | | 3.15E-09 | | 1 | | 0 |
| TP53INP1 | Blood | NA | rs6987752 | | 8:95966531 | | 0.46 | | -0.035 | | 0.004 | | 8.43E-19 | | 145 | | 4892 [In NDUFAF6] |
| USP6NL | Blood | NA | rs968455032 | | 10:11697424 | | NAV | | 0.066 | | 0.009 | | 5.12E-12 | | 61 | | 43671 |
| ECHDC3 | Blood | NA | rs11257290 | | 10:11780324 | | 0.28 | | 0.041 | | 0.005 | | 2.91E-19 | | 115 | | 4041 |
| FRMD4A | Blood | NA | rs1409327 | | 10:13747195 | | 0.41 | | 0.023 | | 0.003 | | 3.40E-13 | | 11 | | 0 |
| SORCS3 | Blood | NA | rs1404786 | | 10:106740404 | | 0.05 | | -0.067 | | 0.007 | | 9.04E-20 | | 223 | | 0 |
| CASP7 | Blood | NA | NAV | | 10:115439640 | | NAV | | -0.072 | | 0.005 | | 8.00E-52 | | 201 | | 0 |
| MS4A2 | Blood | NA | rs514266 | | 11:59877697 | | 0.46 | | -0.094 | | 0.011 | | 5.74E-18 | | 63 | | 14253 |
| MS4A6A | Blood | NA | rs667897 | | 11:59936979 | | 0.47 | | 0.087 | | 0.006 | | 1.91E-50 | | 145 | | 2102 |
| MS4A4A | Blood | NA | rs2162254 | | 11:60039917 | | 0.40 | | 0.109 | | 0.016 | | 2.62E-12 | | 44 | | 8097 |
| PICALM | Blood | NA | rs7131120 | | 11:85690012 | | 0.22 | | -0.051 | | 0.006 | | 7.47E-15 | | 41 | | 0 |
| PSEN1 | Blood | NA | rs214260 | | 14:73662629 | | 0.16 | | 0.076 | | 0.004 | | 1.28E-80 | | 251 | | 0 |
| RIN3 | Blood | NA | rs17783630 | | 14:92955385 | | 0.46 | | 0.037 | | 0.002 | | 3.73E-50 | | 67 | | 24733 [In SLC24A4 ] |
| SLC24A4 | Blood | NA | rs17783630 | | 14:92955385 | | 0.46 | | 0.063 | | 0.004 | | 2.82E-45 | | 73 | | 0 |
| TM2D3 | Blood | NA | rs12907459 | | 15:102225621 | | 0.44 | | 0.062 | | 0.004 | | 3.60E-63 | | 134 | | 33027 [In TARS3] |
| WWOX | Blood | NA | rs7202722 | | 16:78282458 | | 0.40 | | 0.023 | | 0.003 | | 2.60E-14 | | 45 | | 0 |
| PLCG2 | Blood | NA | rs7187863 | | 16:81964977 | | 0.23 | | 0.038 | | 0.004 | | 1.61E-19 | | 68 | | 0 |
| KANSL1 | Blood | NA | rs2732716 | | 17:44323046 | | 0.36 | | -0.087 | | 0.006 | | 4.75E-43 | | 1767 | | 20313 [MAPK8IP1P1(636)] |
| ABCA7 | Blood | NA | rs12462842 | | 19:1100976 | | 0.49 | | 0.017 | | 0.002 | | 2.88E-13 | | 29 | | 35405 [GPX4(2960)] |
| PLD3 | Blood | NA | rs201739636 | | 19:40851678 | | 0.02 | | 0.019 | | 0.003 | | 3.58E-12 | | 8 | | 2685 [In C19orf47] |
| CD33 | Blood | NA | rs200656 | | 19:51724326 | | 0.22 | | 0.038 | | 0.005 | | 1.42E-15 | | 5 | | 3994 |
| SLC24A3 | Blood | NA | rs3827978 | | 20:19281291 | | 0.36 | | 0.043 | | 0.004 | | 1.42E-34 | | 182 | | 0 |
| CASS4 | Blood | NA | rs6014740 | | 20:55045843 | | 0.35 | | 0.040 | | 0.005 | | 1.50E-17 | | 78 | | 11447 [In RTF2] |
| APP | Blood | NA | rs8131895 | | 21:27503527 | | 0.36 | | -0.044 | | 0.004 | | 1.52E-22 | | 126 | | 0 |
| ADAMTS1 | Blood | NA | rs373460567 | | 21:28215827 | | 0.22 | | 0.072 | | 0.006 | | 9.18E-38 | | 27 | | 0 |
| ABCG1 | Blood | NA | rs9976024 | | 21:43641657 | | 0.14 | | -0.030 | | 0.004 | | 2.01E-11 | | 43 | | 0 |
| HLA-DRB5 | Blood | Interferon response(+)/ Anti-bacterial(-) | rs9269047 | | 6:32438783 | | 0.12 | | -7.120 | | 0.335 | | 3.04E-100 | | 9 [all (-)] | | 46337 [HLA-DRB9 (2494)] |
| HLA-DRB5 | Blood | Monocytes/ Macrophages | rs9269047 | | 6:32438783 | | 0.12 | | -11.60 | | 1.030 | | 2.02E-29 | | 1 | | 46337 [HLA-DRB9 (2494)] |
| HLA-DRB5 | Blood | NK cells / CD8+ T-Cells | rs9269047 | | 6:32438783 | | 0.12 | | -7.660 | | 0.994 | | 1.30E-14 | | 1 | | 46337 [HLA-DRB9 (2494)] |
| HLA-DRB1 | Blood | NK cells / CD8+ T-Cells | rs9270928 | | 6:32572461 | | 0.15 | | -4.070 | | 0.377 | | 3.60E-27 | | 287 | | 14836 |
| HLA-DRB1 | Blood | Eosinophils | rs9270994 | | 6:32574250 | | 0.14 | | -2.700 | | 0.415 | | 7.72E-11 | | 42 | | 16625 |
| HLA-DRB1 | Blood | Interferon response(+)/ Anti-bacterial(-) | rs9271147 | | 6:32577385 | | 0.14 | | -5.510 | | 0.250 | | 1.19E-107 | | 346 [260(-)/86(+)] | | 19760 [HLA-DQA1(18571)] |
| HLA-DRB1 | Blood | Monocytes/ Macrophages | rs9271148 | | 6:32577442 | | 0.13 | | -6.110 | | 0.709 | | 6.83E-18 | | 222 | | 19817 [HLA-DQA1(18514)] |
| CR1 | Brain | NA | rs12037841 | | 1:207684192 | | 0.18 | | -0.096 | | 0.007 | | 9.25E-44 | | 64 | | 0 |
| HLA-DRB5 | Brain | NA | rs3117116 | | 6:32367017 | | 0.12 | | -2.780 | | 0.070 | | <1.0E-314 | | 10537 | | 118103 [In TSBP1-AS1] |
| HLA-DRB1 | Brain | NA | rs73399473 | | 6:32538959 | | 0.26 | | -2.050 | | 0.058 | | 8.78E-272 | | 10792 | | 7587 |
| ECHDC3 | Brain | NA | rs866770710 | | 10:11784320 | | 0.0004 | | -0.252 | | 0.018 | | 4.61E-44 | | 45 | | 45 |
| WWOX | Brain | NA | rs12933282 | | 16:78124987 | | 0.45 | | -0.133 | | 0.017 | | 1.13E-15 | | 75 | | 8323 |
| MAPT | Brain | NA | rs2950011 | | 17:43666385 | | 0.23 | | -0.134 | | 0.020 | | 1.65E-11 | | 186 | | 305363 [DND1P1(2090)] |
| OARD1 | Brain | Endothelial Cells | rs17825664 | | 6:405873 | | 0.08 | | -0.812 | | 0.120 | | 1.32E-11 | | 6 | | 40595493 [In IRF4] |
| HLA-DRB5 | Brain | Microglia | rs67987819 | | 6:32497655 | | 0.14 | | -1.900 | | 0.137 | | 9.82E-44 | | 754 | | 0 |
| HLA-DRB5 | Brain | Endothelial Cells | rs67987819 | | 6:32497655 | | 0.14 | | -2.410 | | 0.220 | | 6.32E-28 | | 343 | | 0 |
| HLA-DRB1 | Brain | Microglia | rs72847627 | | 6:32538512 | | 0.28 | | -2.130 | | 0.125 | | 4.15E-65 | | 2305 | | 8034 |
| HLA-DRB1 | Brain | Neurons | rs115480576 | | 6:32538570 | | 0.26 | | -2.210 | | 0.153 | | 2.72E-47 | | 3263 | | 7976 |
| HLA-DRB1 | Brain | Endothelial Cells | rs9269492 | | 6:32542924 | | 0.23 | | -2.250 | | 0.243 | | 2.06E-20 | | 351 | | 3622 |
| HLA-DRB5 | Brain | Neurons | rs9270035 | | 6:32553446 | | 0.14 | | -2.520 | | 0.137 | | 1.46E-75 | | 2540 | | 55382 [In HLA-DRB1] |
| ECHDC3 | Brain | Neurons | rs866770710 | | 10:11784320 | | 0.0004 | | 0.328 | | 0.045 | | 3.13E-13 | | 2 | | 45 |

NA = not applicable; NAV = not available; Chromosome and map position according to GRCh37 assembly; MAF = minor allele frequency; Cell-type specific result rows shaded in gray;

**Table S9**: Significant eGenes located in AD association peaks

| **eGene** | **Tissue** | **Cell-type** | **eSNP+GWAS SNP** | **Position** | **MAF** | **Beta** | **Std Error** | **P-value** | **Nearest AD Gene** |
| --- | --- | --- | --- | --- | --- | --- | --- | --- | --- |
| TRAPPC12 | Blood | NA | rs35067331 | 2:3474085 | 0.29 | 0.021 | 0.003 | 1.23E-10 | TRAPPC12 |
| BIN1 | Blood | NA | rs6733839 | 2:127892810 | 0.38 | -0.040 | 0.003 | 7.42E-38 | BIN1 |
| INPP5D | Blood | NA | rs35349669 | 2:234068476 | 0.46 | 0.025 | 0.003 | 7.88E-14 | INPP5D |
| HLA-DRB1 | Blood | NA | rs9271058 | 6:32575406 | 0.27 | -2.950 | 0.028 | <1.0E-314 | HLA-DRB1 |
| CD2AP | Blood | NA | rs10948363 | 6:47487762 | 0.25 | -0.146 | 0.007 | 2.32E-104 | CD2AP |
| NME8 | Blood | NA | rs2718058 | 7:37841534 | 0.37 | 0.078 | 0.006 | 1.64E-43 | NME8 |
| PILRB | Blood | NA | rs1476679 | 7:100004446 | 0.30 | 0.109 | 0.008 | 1.15E-46 | ZCWPW1 |
| TAS2R60 | Blood | NA | rs11771145 | 7:143110762 | 0.36 | -0.376 | 0.010 | 3.29E-274 | EPHA1 |
| EPHA1 | Blood | NA | rs11771145 | 7:143110762 | 0.36 | -0.029 | 0.004 | 1.90E-14 | EPHA1 |
| PTK2B | Blood | NA | rs28834970 | 8:27195121 | 0.34 | -0.068 | 0.003 | 1.48E-100 | PTK2B |
| TRIM35 | Blood | NA | rs28834970 | 8:27195121 | 0.34 | 0.018 | 0.003 | 3.72E-10 | PTK2B |
| TP53INP1 | Blood | NA | rs1713669 | 8:95958637 | 0.36 | -0.029 | 0.004 | 1.21E-12 | TP53INP1 |
| MADD | Blood | NA | rs3740688 | 11:47380340 | 0.46 | 0.017 | 0.003 | 4.06E-09 | SPI1 |
| MYBPC3 | Blood | NA | rs3740688 | 11:47380340 | 0.46 | -0.020 | 0.002 | 4.47E-18 | SPI1 |
| MS4A6A | Blood | NA | rs983392 | 11:59923508 | 0.41 | 0.072 | 0.006 | 1.22E-33 | MS4A6A |
| MS4A6A | Blood | NA | rs7933202 | 11:59936926 | 0.39 | 0.079 | 0.006 | 4.40E-40 | MS4A2 |
| MS4A4A | Blood | NA | rs1582763 | 11:60021948 | 0.36 | 0.107 | 0.016 | 1.44E-11 | MS4A4A |
| SLC24A4 | Blood | NA | rs10498633 | 14:92926952 | 0.21 | -0.044 | 0.006 | 2.60E-14 | SLC24A4/RIN3 |
| SLC24A4 | Blood | NA | rs12881735 | 14:92932828 | 0.22 | -0.044 | 0.006 | 9.19E-15 | SLC24A4 |
| FAM63B | Blood | NA | rs593742 | 15:59045774 | 0.32 | -0.086 | 0.006 | 8.18E-45 | ADAM10 |
| ARL17A | Blood | NA | rs2732703 | 17:44353222 | 0.21 | 0.147 | 0.023 | 5.95E-11 | MAPT |
| ARL17A | Blood | NA | rs113986870 | 17:44355683 | 0.09 | 0.166 | 0.025 | 2.30E-11 | KANSL1 |
| SUPT4H1 | Blood | NA | rs2632516 | 17:56409089 | 0.47 | -0.036 | 0.004 | 5.14E-22 | BZRAP1 |
| CNN2 | Blood | NA | rs4147929 | 19:1063443 | 0.19 | -0.121 | 0.011 | 7.17E-28 | ABCA7 |
| HMHA1 | Blood | NA | rs4147929 | 19:1063443 | 0.19 | -0.030 | 0.004 | 1.62E-12 | ABCA7 |
| LRRC25 | Blood | NA | rs2303697 | 19:18546678 | 0.35 | 0.043 | 0.003 | 8.37E-57 | ISYNA1 |
| HLA-DRB1 | Blood | Interferon response(+)/ Anti-bacterial(-) | rs9271058 | 6:32575406 | 0.27 | 3.010 | 0.159 | 6.36E-80 | HLA-DRB1 |
| HLA-DRB1 | Blood | NK cells / CD8+ T-Cells | rs9271058 | 6:32575406 | 0.27 | -4.090 | 0.464 | 1.20E-18 | HLA-DRB1 |
| HLA-DRB1 | Blood | Monocytes / Macrophages | rs9271058 | 6:32575406 | 0.27 | -3.540 | 0.497 | 1.06E-12 | HLA-DRB1 |
| CR1 | Brain | NA | rs6656401 | 1:207692049 | 0.17 | -0.096 | 0.007 | 1.05E-43 | CR1 |
| HLA-DRB5 | Brain | NA | rs9271058 | 6:32575406 | 0.27 | -1.690 | 0.081 | 2.28E-106 | HLA-DRB1 |
| HLA-DRB1 | Brain | NA | rs9271058 | 6:32575406 | 0.27 | -1.770 | 0.054 | 1.94E-213 | HLA-DRB1 |
| HLA-DRB5 | Brain | NA | rs9271192 | 6:32578530 | 0.27 | -1.680 | 0.080 | 5.27E-107 | HLA-DRB5 |
| HLA-DRB1 | Brain | NA | rs9271192 | 6:32578530 | 0.27 | -1.760 | 0.054 | 2.82E-213 | HLA-DRB5 |
| KNOP1 | Brain | NA | rs7185636 | 16:19808163 | 0.16 | 0.513 | 0.031 | 2.26E-60 | IQCK |
| LRRC37A2 | Brain | NA | rs2732703 | 17:44353222 | 0.21 | 1.370 | 0.053 | 4.13E-150 | MAPT |
| ARL17A | Brain | NA | rs113986870 | 17:44355683 | 0.09 | 1.260 | 0.047 | 4.96E-12 | KANSL1 |
| LRRC37A2 | Brain | NA | rs113986870 | 17:44355683 | 0.09 | -0.326 | 0.068 | 1.98E-76 | KANSL1 |
| HLA-DRB1 | Brain | Neurons | rs9271058 | 6:32575406 | 0.27 | -1.400 | 0.135 | 2.37E-34 | HLA-DRB1 |
| HLA-DRB5 | Brain | Neurons | rs9271058 | 6:32575406 | 0.27 | -1.650 | 0.201 | 1.24E-14 | HLA-DRB1 |
| HLA-DRB1 | Brain | Microglia | rs9271058 | 6:32575406 | 0.27 | -1.550 | 0.111 | 1.80E-36 | HLA-DRB1 |
| HLA-DRB1 | Brain | Endothelial Cells | rs9271192 | 6:32578530 | 0.27 | -1.400 | 0.245 | 2.96E-12 | HLA-DRB5 |
| HLA-DRB1 | Brain | Neurons | rs9271192 | 6:32578530 | 0.27 | -1.650 | 0.135 | 2.37E-34 | HLA-DRB5 |
| HLA-DRB5 | Brain | Neurons | rs9271192 | 6:32578530 | 0.27 | -1.550 | 0.201 | 1.24E-14 | HLA-DRB5 |
| HLA-DRB1 | Brain | Microglia | rs9271192 | 6:32578530 | 0.27 | -1.710 | 0.110 | 4.17E-37 | HLA-DRB5 |
| LRRC37A2 | Brain | Microglia | rs2732703 | 17:44353222 | 0.21 | 1.520 | 0.147 | 7.65E-24 | MAPT |
| LRRC37A2 | Brain | Neurons | rs2732703 | 17:44353222 | 0.21 | 1.480 | 0.140 | 1.84E-27 | MAPT |
| LRRC37A2 | Brain | Endothelial Cells | rs2732703 | 17:44353222 | 0.21 | 1.750 | 0.233 | 5.88E-14 | MAPT |
| LRRC37A2 | Brain | Neurons | rs113986870 | 17:44355683 | 0.09 | 1.530 | 0.184 | 2.77E-14 | KANSL1 |
| LRRC37A2 | Brain | Microglia | rs113986870 | 17:44355683 | 0.09 | 1.400 | 0.195 | 4.29E-15 | KANSL1 |
| Known AD gene |  |  |  |  |  |  |  |  |  |

NA = not applicable; Chromosome and map position in base pairs; MAF = minor allele frequency; Cell-type specific result rows shaded in gray;

**Table S10:** Cell-type distribution of significant ct-eQTLs

| **Tissue / cell-type** | **# significant**  **ct-eQTLs** | **% significant**  **ct-eQTLs** | **Reference % in tissue^1^** |
| --- | --- | --- | --- |
| **Blood** |  |  |  |
| Neutrophils.1 | 1,007 | 3.3 | 53.8±6.1 of leukocytes^2^ |
| Neutrophils.2 | 50 | 2.4 | 53.8±6.1 of leukocytes |
| CD4+ T-cells | 562 | 1.8 | 14.6±3.5 of leukocytes |
| NK cells / CD8+T-cells | 2,611 | 8.6 | NK cells=4.4±2.4 of leukocytes  CD8+ T cells=6.8±1.3 of leukocytes |
| Erythrocytes | 2,130 | 7.0 | 93-96% of blood cells |
| Monocytes/macrophages | 3,234 | 10.6 | 8.4±1.3 of leukocytes |
| Interferon response /  Anti-bacterial cells | 19,331 | 63.6 | NA |
| B-cells | 89 | 0.3 | 5.2±2.3 of leukocytes |
| Eosinophils | 735 | 2.4 | 3.2±1.6 of leukocytes |
| Unknown | 646 | 2.1 | NA |
| **Brain** |  |  |  |
| Endothelial cells | 10,597 | 20.7 | ~2:1 glia^3^ to endothelial cells |
| Neurons | 18,930 | 37.0 | 1:1 ratio of glia to neurons |
| Microglia | 15,560 | 30.4 | 10% of glial cells |
| Astrocytes | 1,040 | 2.0 | 19–40% of glial cells |
| Oligodendrocytes | 4,971 | 9.7 | 45–75% of glial cells |

^1^References in blood [37, 38] and in brain [39, 40]

^2^ Leukocytes: 0.1-0.2% of blood cells

^3^ Glial cells include microglia, oligodendrocytes and astrocytes

**Table S11:** Gene-set enrichment analysis including eGenes of significant ct-eQTLs in myeloid cell lineages

| **PANTHER Pathway** | | [**#**](http://pantherdb.org/tools/compareToRefList.jsp?sortOrder=2&sortList=Homo%20sapiens) **Genes Annotated to Pathway** | | **# Genes in Network** | | **Expected P value *** | **Fold Enrichment** | | **Direction** | | **Unadjusted**  **P-value** | | **FDR** |
| --- | --- | --- | --- | --- | --- | --- | --- | --- | --- | --- | --- | --- | --- |
| Alzheimer disease-amyloid secretase pathway | | 67 | | 5 | | 0.41 | 12.06 | | + | | 8.18E-05 | | 1.34E-02 |
| Oxytocin receptor mediated signaling pathway | | 58 | | 4 | | 0.36 | 11.15 | | + | | 5.78E-04 | | 4.74E-02 |
| Thyrotropin-releasing hormone receptor signaling pathway | | 60 | | 4 | | 0.37 | 10.78 | | + | | 6.52E-04 | | 3.56E-02 |
| 5HT2 type receptor mediated signaling pathway | | 67 | | 4 | | 0.41 | 9.65 | | + | | 9.64E-04 | | 3.95E-02 |

PANTHER = Protein ANalysis THrough Evolutionary Relationships) Classification System; * Expected probability of observing at least x number of genes out of the total n genes annotated to the pathway; FDR = false discovery rate

**Fig. S1:** Study design

All protein-coding genes

**Genotype data**

Filtered to

- MAF ≥ 0.05
- Imputation quality R^2^ ≥ 0.3

**Cis- eQTL mapping for SNPS ± 1 Mb around each gene**

**Genome-wide**

**Interaction analyses of all eQTL-proxy gene combinations**

**= cell-type specific eQTLs**

**Blood and brain**

**cell-type proxy genes**

**Fig. S2:** Regional plots for colocalized AD GWAS/lead eQTL variant pairs. **A)** rs10948363 *CD2AP*/ rs4711880 *CD2AP;* **B)** rs28834970 *PTK2B*/ rs6557994 *PTK2B;* **C)** rs9331896 *CLU*/ rs6557994 *PTK2B;* **D)** rs10838725 *CELF1*/ rs35233100 *MADD;* **E)** rs983392 *MS4A6A*/ rs11230563 *CD6;* **F)** rs429358 *APOE*/ rs74253343 *RELB*.

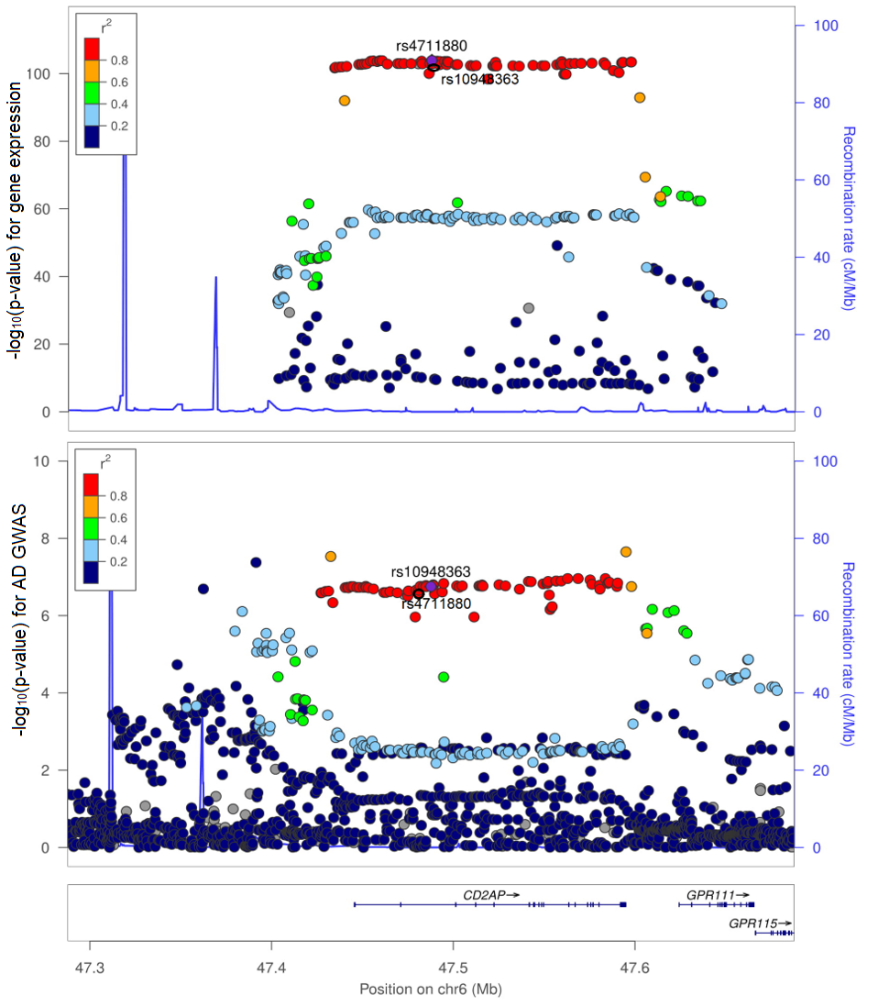

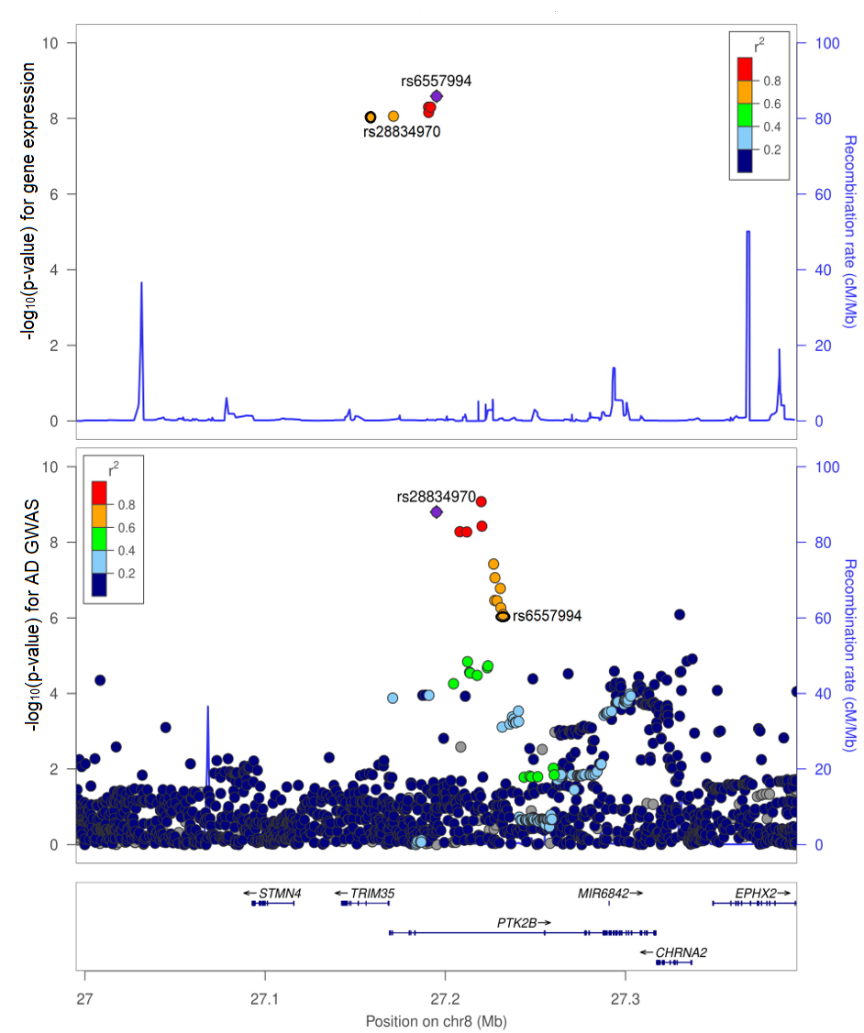

**A)**

**B)**

**D)**

**C)**

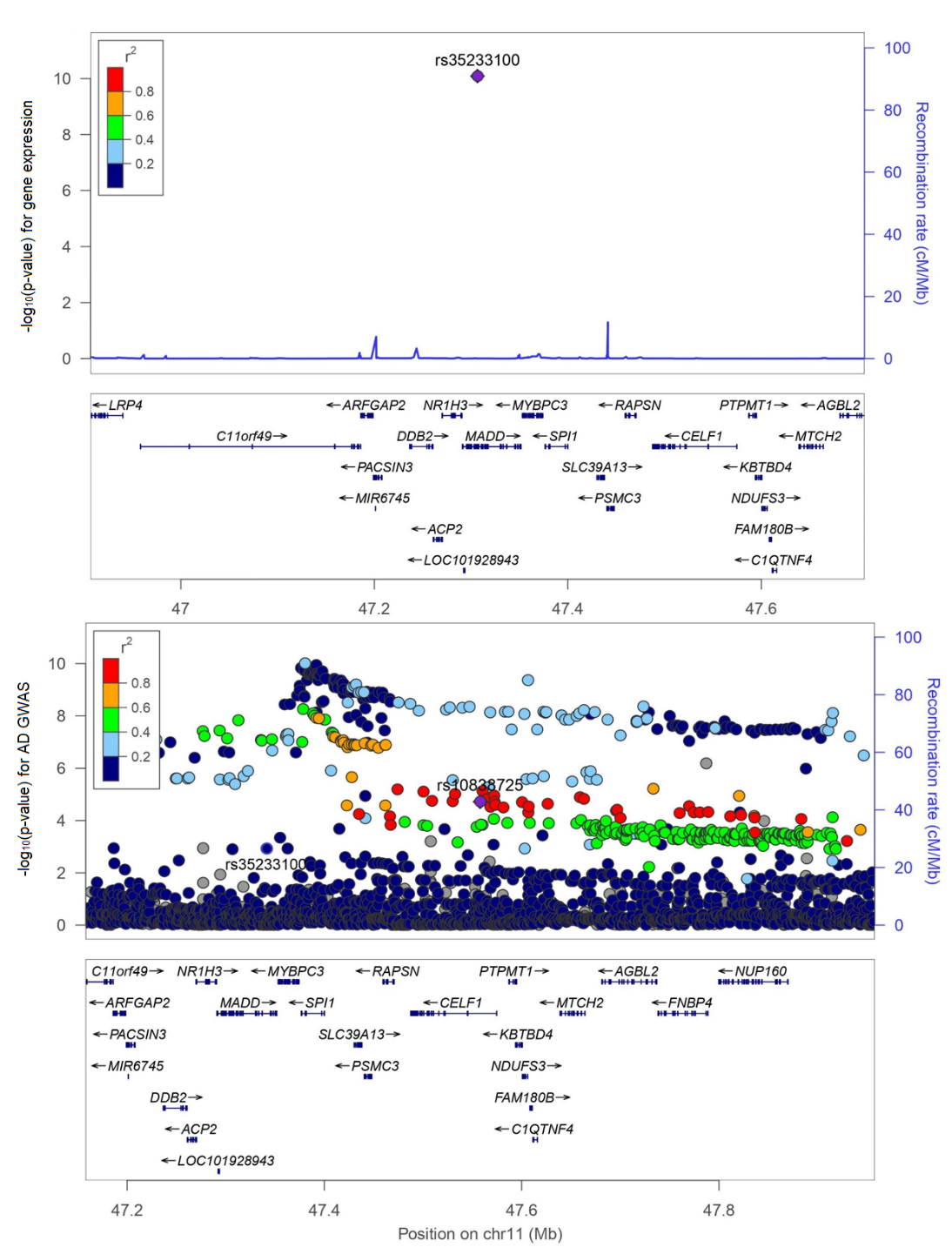

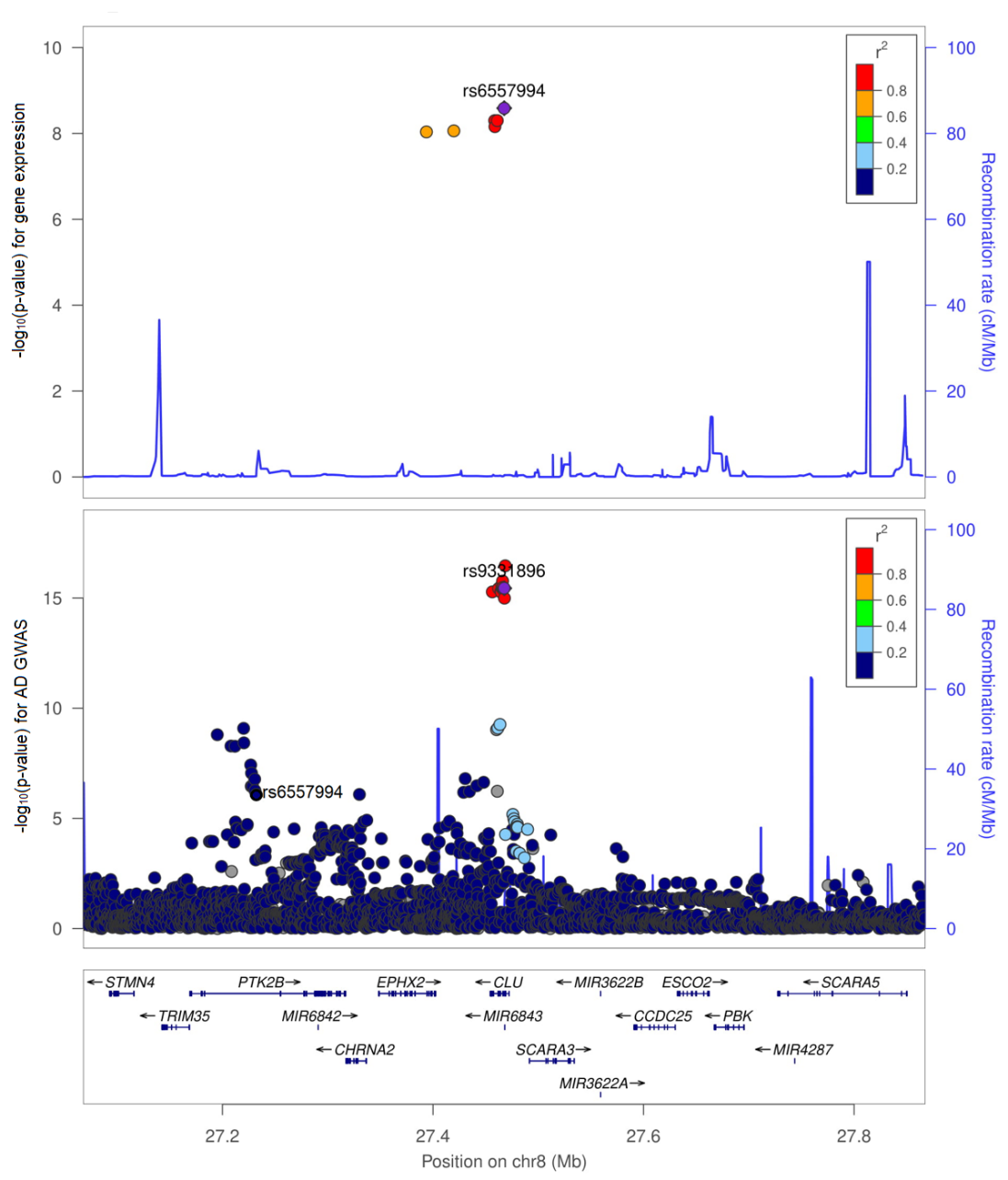

**E)**

**F)**

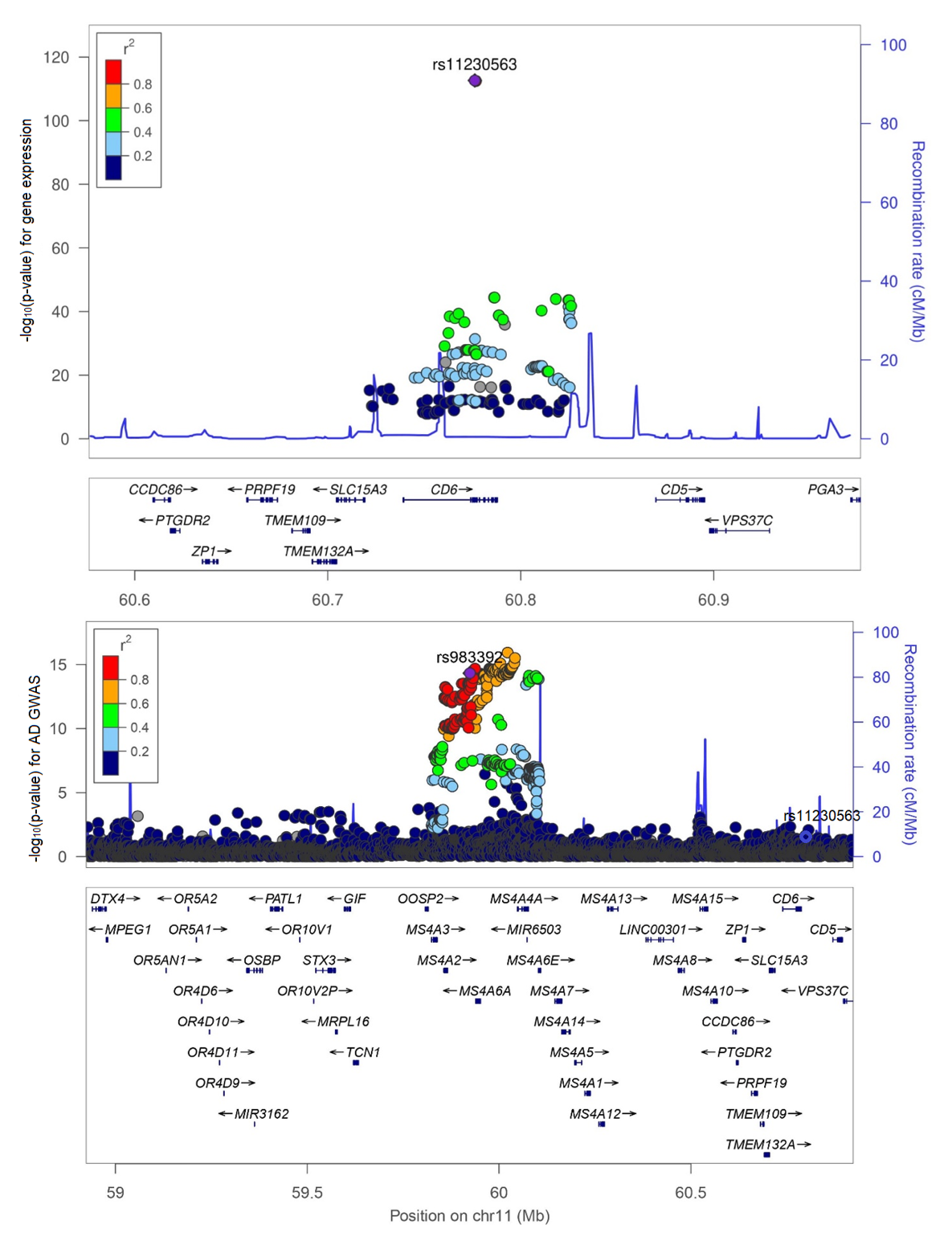

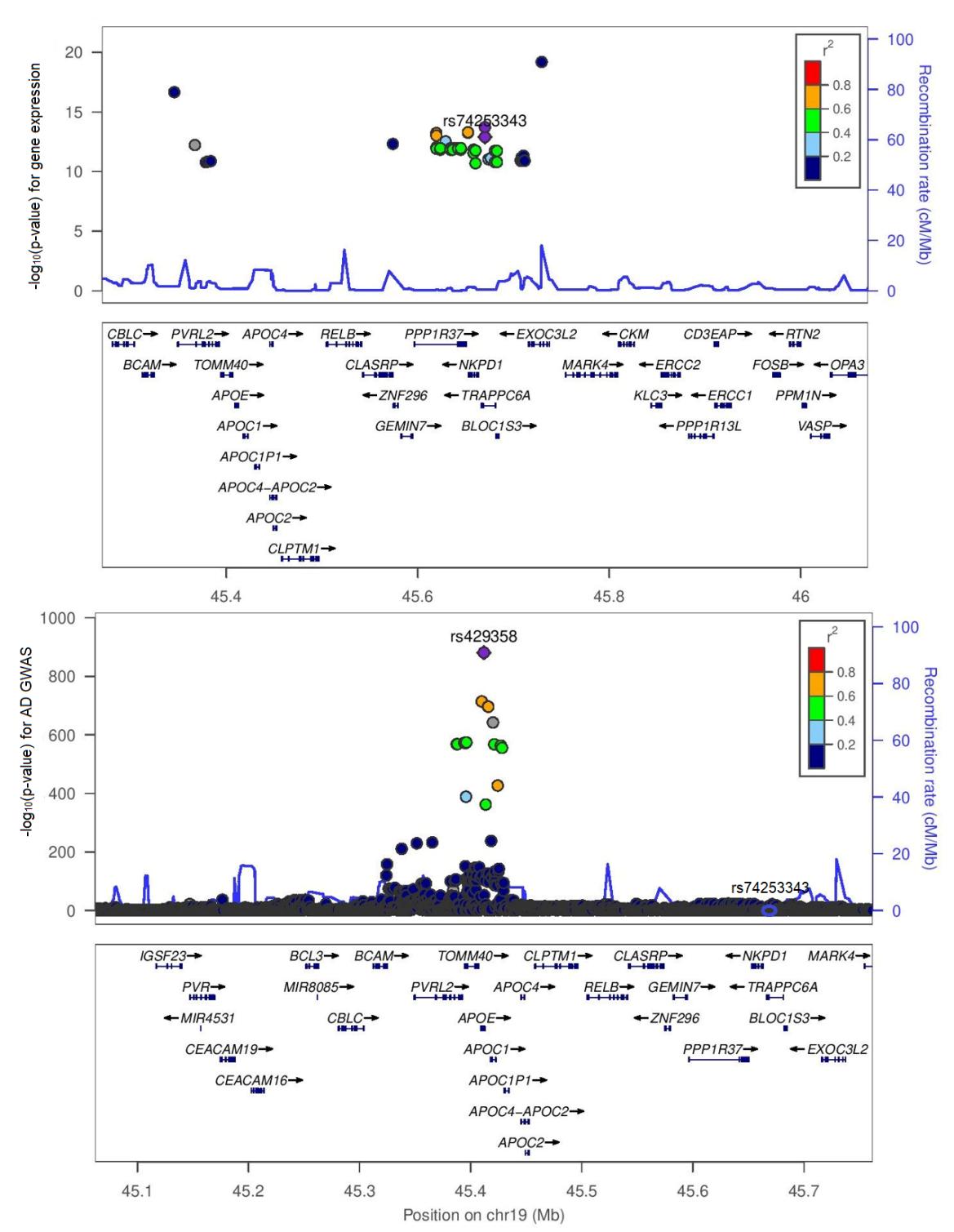
